# From warehouse to ward: Applying implementation research methods to the device identification, qualification, distribution, and management process within the NEST360 alliance

**DOI:** 10.1101/2025.05.28.25328509

**Authors:** Kylie Dougherty, Rebecca P. Kirby, Katerina Claud, Kara M. Palamountain, Elizabeth Asma, Millicent Alooh, Danica Kumara, Vince Gate, Nicki Bisceglia, Ali Khalid, Vincent Otieno Ochieng, George Banda, Hope David Peter, Elizabeth Ngowi, Charles Osuagwu, Rebecca Richards-Kortum, Z. Maria Oden, Christine Bohne, Lisa R Hirschhorn

**Author notes:** Corresponding Author, Kylie Dougherty, Northwestern University Feinberg School of Medicine 625 Michigan Ave., Chicago, IL 60611.

## Abstract

**Background:** Preventing newborn deaths is possible with the right medical devices. However, in many countries, devices for small and sick newborn care (SSNC) are either unavailable, not fit for the environment, or broken. Newborn Essential Solutions and Technologies (NEST360) is a multicountry interdisciplinary alliance aimed at decreasing neonatal mortality in Kenya, Malawi, Nigeria, and Tanzania. The technology qualification, distribution and management teams in NEST360 work to ensure appropriate devices are available and functional at facilities. Their work involves identifying which technologies are needed, sourcing devices that fit those needs, testing those devices’ functional ability under varied conditions, establishing a reliable supply chain, training facility staff in use, and maintaining the devices once installed through trained biomedical technicians. We applied implementation research (IR) to understand context, describe strategy selection, and implementation outcomes of the work designed to ensure the consistent availability of functional devices for SSNC.

**Methods:** Between March and July 2024, we conducted in-depth interviews with NEST360 team members via Zoom, and reviewed quantitative programmatic data, including device functionality reports. We applied deductive content analysis for interviews and descriptive statistics for quantitative data. Results were used to develop an implementation research logic model (IRLM) using NEST360 and UNICEF’s SSNC Implementation Toolkit for contextual factors and RE-AIM for implementation outcomes.

**Results:** We identified 40 contextual factors, 78% being barriers. Twenty-one strategies were implemented to address barriers to device qualification, distribution, and management efforts, including engaging stakeholders and conducting ongoing trainings. Notable implementation outcomes included **Reach** with 29 devices in 12 product categories qualified, and all 66 facilities received NEST-qualified devices**, Effectiveness**, in 2024, an average of 87% of all newborn care devices were functional, including those provided by NEST360 and those sourced through existing channels, **Adoption** with over 2,476 devices installed at NEST360 sites in 2023**. Acceptability** was also high with country-level biomedical technicians reporting positive facility-level experiences using the devices.

**Conclusions:** NEST360 approach to ensuring appropriate and functioning equipment for SSNC was successful through multiple strategies to address multilevel barriers. The use of IR facilitated understanding of how strategies addressed context and where change is needed. These results will be used in plans for scale-up and dissemination.

**Contributions to the literature:**

- This study utilizes implementation research (IR) methodologies to examine the contextual factors, strategies, and outcomes of maintaining functional medical devices for small and sick newborn care (SSNC) in four countries. By developing an implementation research logic model (IRLM), this study provides a structured approach to understanding how device access and sustainability can be improved in resource-limited settings.
- The study identified 40 contextual factors in multiple areas and levels influencing the availability of functional devices, including human resource constraints, governance issues, financial barriers, and infrastructure challenges. It maps these barriers and facilitators to 21 targeted strategies that align with the Expert Recommendations for Implementation Research (ERIC) framework, providing a blueprint for overcoming diverse challenges.
- The study explores long-term sustainability of the NEST360-supported work by identifying key areas for improvement, such as financing for spare parts, strengthening preventive maintenance practices, and advocating for policy reforms.

## 1. Background

Neonatal mortality is a significant challenge globally, and accounts for almost half of under-five deaths in 2022 (1). To achieve the 2030 Sustainable Development Goal 3.2 of ending preventable deaths of newborns, the United Nations led a global collaboration to develop Every Woman Every Child Everywhere. This plan outlines the steps for reducing newborn mortality and emphasizes the need to scale up small and sick newborn care (SSNC) in level 2+ neonatal units including in low and middle income countries (2). Level 2+ units provide specialized care for newborns, including respiratory support for preterm infants and other essential interventions. Preventing newborn death and providing high-quality SSNC requires facilities to have functional medical devices such as radiant warmers, continuous positive airway pressure (CPAP) machines, phototherapy machines, and pulse oximeters. However, many countries and facilities lack the consistent availability of these devices and thus the quality of care that they can provide neonates (3, 4). Previous research has found that poorly managed and coordinated programs providing medical equipment can lead to the development of device graveyards, where non-functional or inappropriate devices accumulate in healthcare facilities (5). One study estimates that nearly 40% of medical equipment in developing countries is non-functional due to maintenance failures, a shortage of spare parts and accessories, and a lack of trained biomedical engineers, discrepancies between device specification and facility capabilities, and insufficient clinician training (6, 7).

Newborn Essential Solutions and Technologies (<u>NEST360</u>) is a multi-country alliance committed to reducing deaths among small and sick newborns in level 2+ SSNC units across four African countries: Kenya, Malawi, Nigeria, and Tanzania. NEST360, in partnership with African governments, supports the implementation of a comprehensive care package through various strategies including data for action, quality improvement (QI), and training. One strategy is designed to ensure access to appropriate and affordable devices, such as pulse oximeters and radiant warmers; providing training for clinicians in SSNC and biomedical engineers and technicians (BMETs) in correct device use, maintenance, troubleshooting, and repair (5, 8–11). While NEST360 supports this work by identifying and qualifying essential technologies, it does not manufacture devices; however, within the broader NEST360 alliance, Rice360 and 3^rd^ Stone Technologies-both member organization-have produced three qualified devices, following the same rigorous standards applied to all devices evaluated by NEST360. In Phase 1 (2019-2023), NEST360 has supported 68 SSNC units in four countries in sub-Saharan Africa (Kenya, Malawi, Nigeria, Tanzania). However, even with this support, some of NEST360’s facilities have experienced difficulty maintaining functional devices. A previous publication from the alliance explored pre-implementation facility-level readiness to provide CPAP, and their analysis found that readiness ranged from 50% of facilities in Malawi, 46% of sites in Kenya, 33% of sites in Tanzania, and 21% of sites in Nigeria, which identified a need to explore the contextual factors and strategies that influenced NEST360’s device implementation and maintenance efforts (12).

Implementation Research (IR) methods can help understand the link between contextual factors, chosen strategies and outcomes of interventions to both help improve implementation and create generalizable knowledge. This knowledge can help to provide a systematic approach that can be scaled to larger audiences, help identify positive contextual factors and implementation strategies that are critical to drive change (13, 14). In this paper, we used IR methods and frameworks to develop an implementation research logic model (IRLM) to understand contextual factors and strategies used to address them and associated implementation outcomes of NEST360’s work to ensure access to appropriate, affordable and functional devices, as well as the identification and procurement of those devices. The results will support NEST360 to identify strategies needing adaptations to promote implementation success. These findings can also inform the scaling of the NEST360 program to other sites and inform other programs working to improve the availability of functional devices for SSNC or other clinical needs.

## 2. Methods

### 2.1 Settings

The study used data from all 66 facilities in Kenya (n=13), Malawi (n=37), Nigeria (n=9), and Tanzania (n=7) that receive support from NEST360. The sample included all sites supported by NEST360 across participating countries, with a higher number from Malawi due to its nationwide rollout of the NEST360 program. Level 2+ units provide specialized care for newborns, including respiratory support through CPAP and oxygen, and other essential interventions. These facilities were selected because they met the service readiness criteria established by the NEST360 alliance, example criteria included the presence of a separate room for neonatal care, backup power for the neonatal unit, and standard forms and registers used in the neonatal unit (11).

### 2.2 Guiding frameworks

This retrospective study leveraged several IR frameworks to explore NEST360’s work aimed at maintaining functional devices at their supported facilities. The NEST360/ United Nations Children’s Fund (UNICEF) SSNC core components framework, as adapted by the Newborn Toolkit community, was used to identify and categorize contextual factors affecting the consistent availability of functional devices (Figure 1) (15, 16). The NEST360 Newborn Toolkit Community is a global network of over 300 implementers—policymakers, healthcare professionals, engineers, and researchers—collaborating to improve care for small and sick newborns (17). This community co-developed the open-access Implementation Toolkit, which provides practical resources to support country-led efforts in delivering high-quality newborn care (17). This framework was chosen because it outlines the components of health systems that influence SSNC delivery, which are critical for facilities to be able to maintain functional devices and use them appropriately (15, 16).

**Figure 1.**
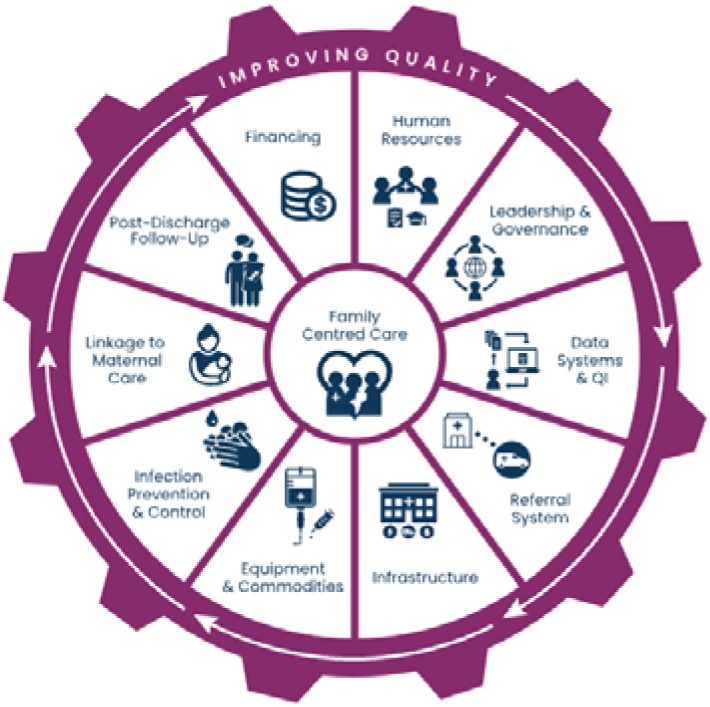
**Small and sick newborn care core components framework adapted by the Newborn Toolkit community(16)**

We used the Expert Recommendations for Implementation Research (ERIC) framework to categorize implementation strategies (18). We used RE-AIM outcomes to measure the outcomes of *reach, effectiveness, adoption, implementation (including acceptability, adaptation, feasibility, fidelity), and maintenance* of the work (Table 1) (19). Implementation outcomes measured multi-level outcomes including facility-level, country-level, and alliance-level. We used the data collected and reviewed to develop an IRLM. We chose to develop a logic model because it is a simple way to map contextual factors to the strategies targeting them as well as the intended implementation outcomes (20).

**Table 1.** RE-AIM outcomes and how they were operationalized in this study (19)

| Dimension | Operationalized |
| --- | --- |
| Reach | • Number of facilities that received technology from NEST360 by the end of Phase 1 (2023) |
|  | • Number of devices (specific brand and type) that have received NEST360 qualification by the end of Phase 1 (2023) |
|  | • Number and percentage of categories that have at least one qualified device approved by the end of Phase 1 (2023) |
| Effectiveness | • Percentage of facilities with enough CPAP devices to cover all eligible babies by the end of Phase 1 (for most facilities this was measured in 12/2023, but some it was measured in 9/2023) |
|  | • Percentage of facilities with enough phototherapy devices to cover all eligible babies by 12/2023 |
|  | Percentage of all newborn care devices that were functional, includes those provided by NEST360 and those sourced through existing channels (2024) |
| Adoption | Number of NEST360 qualified devices installed at NEST360 facilities by the end of Phase 1 (2023) |
| **Implementation** |  |
| *Acceptability* | Responses from device team members on acceptability of the identification, qualification, distribution, and management process (Interviews 5/24-7/24) |
| *Adaptations* | Changes that occurred to the device identification, qualification, distribution and management process by the end of Phase 1 (2023) |
| *Feasibility* | Percentage of facilities with a BMET available during day or night shift by the end of Phase 1 (for most facilities this is 12/2023, but some it is 9/2023) |
|  | Responses from device team members on distributors' ability to get NEST360 qualified devices into countries and installed at facilities by the end of Phase 1 (2023) |
| *Fidelity* | Percentage of facilities performing monthly PPM for 5 of the 6 reported categories (glucometer, pulse ox, CPAP, O2 concentrator, radiant warmer, phototherapy) (Comparison 2/22-12/22 and 2/23-12/23) |
|  | Number of NEST360 qualified devices that underwent the full identification and qualification process by the end of Phase 1 (2023) |
| **Maintenance** | Average number of facilities performing monthly PPM for 5 of the 6 reported categories (glucometer, pulse ox, CPAP, O2 concentrator, radiant warmer, phototherapy) (1/24-6/24) |
|  | Responses from device team members about areas where the device teams have already, are beginning to, or should consider focusing their attention to ensure sustainability of their work (Interviews conducted 5/24-7/24) |

*Note.* CPAP (Continuous positive airway pressure); BMET (Biomedical engineers and technicians); PPM (Planned preventative maintenance)

We used the Standards for Reporting Implementation Studies (StaRI) checklist (See supplementary material 1) to guide the reporting of our implementation study (21).

### 2.3 Data Sources and collection

#### Quantitative data

To measure quantitative implementation outcomes, we used routine programmatic data that are available within NEST360’s data dashboard (NEST-Implementation Tracker (NEST-IT)), and device monitoring programs (22). NEST-IT presents interdisciplinary indicators at multiple levels (facility, country, alliance) (22). The data in NEST-IT comes from facilities’ routine reports that document human resources, infrastructure, the incidence of planned preventative maintenance (PPM) occurring for devices, ratios of devices to newborns, receipt of essential care for SSNC including those requiring functioning equipment (ex. CPAP, phototherapy) and clinical and coverage indicators that are extracted from NEST360’s Neonatal Inpatient Dataset (NID), which is a data tool used in routine information systems to inform inpatient SSNC quality (8, 22). Asset tracking and ticketing programs report device inventory and installation tracking, including functional status and location, and device issues or downtime which are tracked through tickets and activity scheduling (22).

#### Qualitative data

We also conducted key informant interviews (KIIs) with NEST360 team members. These interviews provided details and context for the quantitative implementation outcomes. The interviews focused on the steps taken from a device being identified to the point where it is maintained at NEST360-supported facilities. The interviews explored barriers and facilitators (factors that enhance or impede a facility’s ability to maintain functional devices), actions and strategies NEST360 used to enable this work, and provided additional context for understanding implementation outcomes. Participants were purposively sampled from the different teams within NEST360 that work on newborn devices (device qualification, management, and distribution teams). The device qualification team works to identify technology that could be used to support SSNC using an 8-step evidence-based technology review process (5). The health technology management team supports both device management and distribution. The device management work assists with the installation of the devices at the facilities as well as providing training and mentorship for clinicians and BMETs on using and maintaining the devices, and documentation of device related data. Furthermore, device management supports the procurement and availability of accessories, consumables, and spare parts (ACS). Individuals involved in device distribution assist with ordering the devices for NEST-supported facilities, ensuring the devices have the appropriate paperwork to be imported into the country, and facilitating the shipment of devices from the manufacturer to the facilities. To recruit participants, a team member reached out to the possible participants via email to provide a brief overview of the study, describe the KII, and request participation. All individuals recruited were told that their participation was completely voluntary. To participate in the KIIs, individuals had to be at least 18 years old, speak English, work directly within the NEST360 alliance, and have job tasks that directly involved SSNC devices.

We used an iteratively refined interview guide with open-ended questions to explore contextual factors using the SSNC framework, strategies, and gain a deeper understanding of preliminary implementation outcomes (see supplemental material 2). The interviews were conducted in English via Zoom, with both video recordings and transcripts generated using Zoom’s built-in features (23). Interviewers were led by KD and lasted between 35-60 minutes. During the interviews, we conducted member checks by summarizing the key concepts and ideas expressed by participants and asked for their confirmation to ensure we accurately represented their perspectives (24).

### 2.5 Data management

For the quantitative data the country specific data was stored and managed by each country on a secure server that met their country’s specific requirements for health data. The authorship team only had access to de-identified data transferred to a central pooled database for analysis via the NEST360 program data dashboard, NEST-IT. For the qualitative data all interviewees were identified by a participant number. The participant numbers were linked to their names and stored behind an encrypted, password-protected site. Transcripts generated by Zoom were checked for accuracy against the recordings and deidentified before sharing them with other research team members. We used Dedoose to manage, code, and analyze all qualitative data (25).

### 2.6 Analysis

We used a qualitative descriptive methodology and performed deductive directed content analysis (26). This analysis was guided by a codebook developed by KD and KC (see supplemental material 3), based on the constructs of the SSNC core components, and the key components of the Stanford Lightning Report (successes, challenges, and suggestions) (16, 27). KC independently coded the transcripts, identifying contextual factors (barriers, facilitators), strategies that aligned with the SSNC core components, successes of these activities, challenges, and participant suggestions, and documented the process (16, 27). KD reviewed the documentation and worked with KC to refine the codes, ensuring the confirmability of the coding decisions. The contextual factors were then categorized as either facilitators (i), barriers (!), or both (i/!) based on whether they enabled the availability of functional devices (i), or prevented it (!), or could both enable and prevent the availability of functional devices (i/!). Strategies extracted from the transcripts that addressed SSNC core components were then mapped to the ERIC strategies that were most closely aligned (18). Any adaptations to strategies from the original strategies, as well as participants responses related to the acceptability of NEST360’s device activities were also extracted from the transcripts. The extracted data on successes, challenges, and recommendations were synthesized into Stanford Lightning Reports, a qualitative method designed to generate concise summaries for the rapid reporting of emerging qualitative findings (27).

Facility- and country-level programmatic data and qualitative findings were used to measure implementation outcomes. We used descriptive statistics for quantitative data.

The contextual factors, strategies, and implementation outcomes were combined into an IRLM. Multiple iterations of the complete IRLM were shared via email with NEST360’s device qualification, distribution, and management team members to ensure consensus and completeness.

## Results

A total of twelve KIIs were conducted, all individuals asked to participate agreed to be a part of the study. Table 2 provides demographic details for the participants.

**Table 2.**
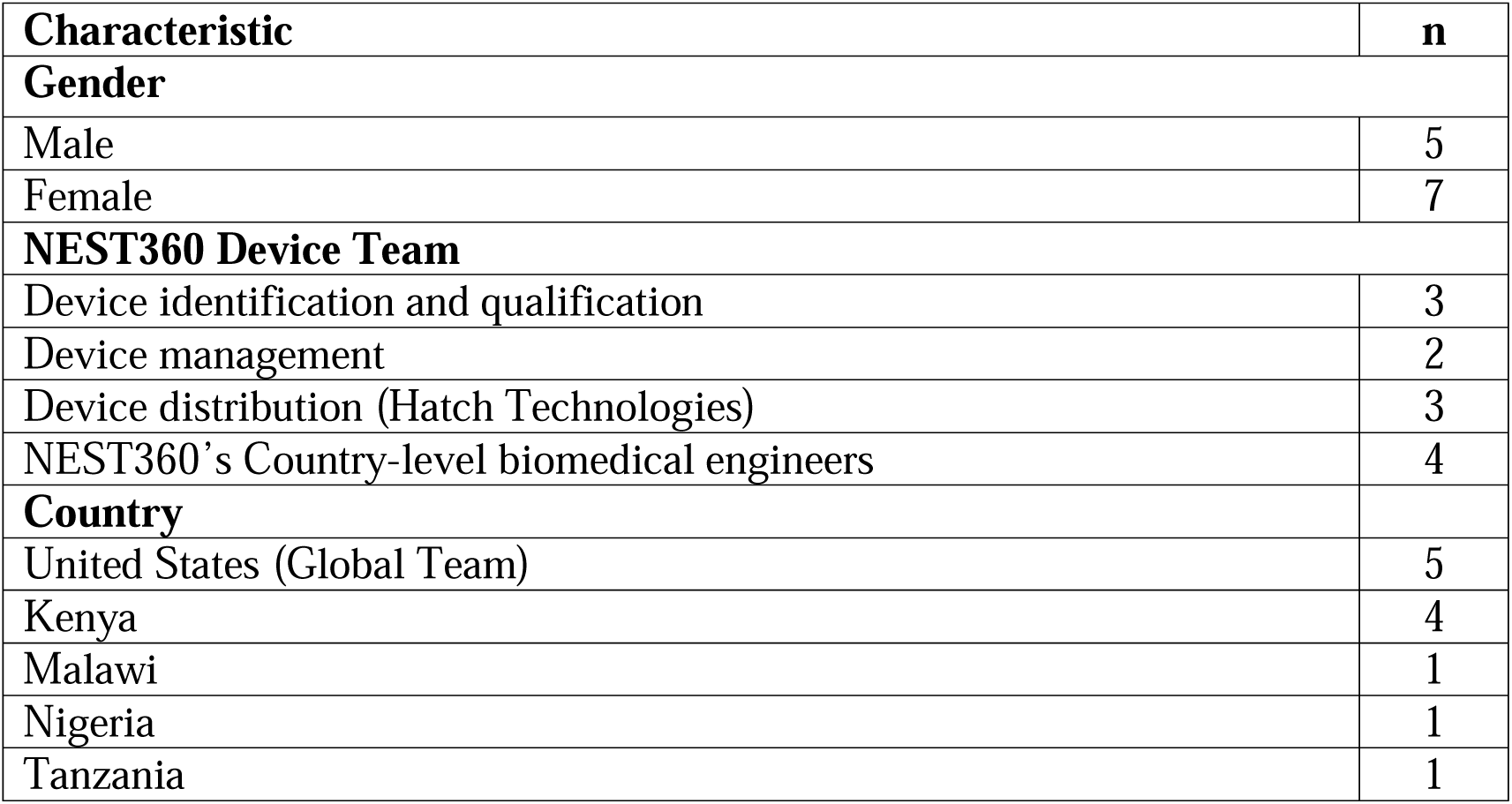
Demographic characteristics of key informant interview participants.

### 3.1 Contextual factors

We identified 40 contextual factors, 78% (31 factors) were barriers and 20% (8 factors) were facilitators, 2% (1 factor) was seen as both a barrier and facilitator (Table 3). These contextual factors influenced the facility, country, and alliance’s ability to ensure the consistent availability of functional devices.

**Table 3.** Contextual factors for the implementation of in NEST360 organized by the SSNC core components.

| Domain | Contextual Factor | Barrier (↓),<br>Facilitator (↑), Both<br>(↑/↓) |
| --- | --- | --- |
| **Human Resources** | BMETs unavailable | ↓ |
|  | Staff turnover | ↓ |
|  | BMETs lack skill with maintaining and refurbishing devices | ↓ |
|  | Clinicians lack skill with using the devices properly | ↓ |
|  | Lack of collaboration between BMETs and clinicians | ↓ |
|  | Inconsistent BMET training | ↓ |
|  | Motivated staff | ↑ |
| **Equipment & Commodities** | Expensive equipment for SSNC | ↓ |
|  | Expensive accessories, consumables, and spare parts | ↓ |
|  | No procurement system | ↓ |
|  | Missing tools for BMETs to perform maintenance | ↓ |
|  | Stockouts | ↓ |
|  | SSNC devices not designed for low-resource/harsh environments | ↓ |
|  | Complex and frequent maintenance needs for devices | ↓ |
| **Financing** | NEST360 grant funding | ↑ |
|  | Lack of government financing | ↓ |
|  | Lack of long-term funding | ↓ |
|  | Lack of budget allocations for spare parts and accessories | ↓ |
| **Service Delivery** | Local distributors | ↑/↓ |
|  | PPM not occurring | ↓ |
| **Leadership & Governance** | Global and national focus on SSNC and NMR | ↑ |
|  | BMETs not prioritized at the national or facility level | ↓ |
|  | Difficult device import regulations and clearance requirements | ↓ |
|  | Exclusive single distributor contracts for certain devices | ↓ |
|  | Shipping and import delays | ↓ |
|  | WHO guidelines on priority medical devices for SSNC | ↑ |
|  | Collaboration with UNICEF | ↑ |
|  | NEST360 interdisciplinary expertise | ↑ |
|  | Device catalogs, technology landscape documents, durability testing documents | ↑ |
|  | Competing interests/ priorities between global funders | ↓ |
|  | No single cross-country regulatory authority for newborn devices in Africa | ↓ |
| **Data Systems & QI** | Facilities tracking clinical outcomes | ↑ |
|  | Infrequent data use | ↓ |
|  | No system for monitoring BMET-specific indicators | ↓ |
|  | Infrequent BMET QI projects | ↓ |
| **Infrastructure** | Unavailable workspaces for BMETs | ↓ |
|  | No space to store all SSNC devices | ↓ |
|  | Power surges and outages | ↓ |
|  | Lack of HVAC available at the facility | ↓ |
|  | Harsh environmental conditions | ↓ |

*Biomedical engineers and technicians (BMETs); Small and sick newborn care (SSNC); Planned preventative maintenance (PPM); Neonatal mortality rate (NMR); World health organization (WHO); United nations children's fund (UNICEF); Quality improvement (QI); Heating, ventilation, and air conditioning (HVAC)*

In *Human Resources*, there were several barriers that NEST360 needed to address through various strategies, such as staff availability and skill. This included both clinicians and BMETs since trained staff would be needed to use the devices with newborns correctly, and BMETs critical to providing preventative maintenance and fixing broken devices. While there were several barriers surrounding human resources, one facilitator that NEST360 leveraged was the highly motivated staff, both clinicians and BMETs, who engaged in NEST360’s implementation efforts.

Prior to NEST360’s involvement, *equipment and ACS,* had several barriers that significantly impacted facilities’ abilities to maintain functional SSNC devices. These barriers included devices frequently needing spare parts for repair, devices that may be available but were not designed for use in challenging environments that experience high temperatures or power surges. These devices, often, required frequent or complex maintenance, and originally many BMETs did not have the required tools such as multimeters, analyzers, wrenches and screwdrivers to perform maintenance and refurbishment at the facilities.

*Financing* barriers persisted because many facilities did not have budgets allocating resources for BMETs or spare parts, and BMETs were often not prioritized when thinking about government financing. One facilitator though was the grant funding that NEST360 provided to support their work.

In *service delivery,* PPM showed improvement from baseline with the average number of facilities performing monthly PPM for five of six reported devices increasing from 58.6% in 2022 to 70.8% in 2023. Additionally, in some of the countries where NEST360 works there were local distributors. This was both a facilitator and barrier to NEST360’s work because these local distributors understood their context, and the manufacturers that they had worked with previously. However, in some cases, it was also a barrier because these local distributors might have exclusive contracts with manufacturers and then NEST360 distributors were unable to provide that product to local facilities.

*Leadership and governance* had the most facilitators identified. This is for many reasons, such as a current global focus on SSNC and the prevention of newborn mortality. Additionally, the World Health Organization’s published guidelines defining the priority medical devices for providing SSNC, and device catalogs, landscaping documents, and durability testing guidelines the availability and NEST360 was able to use these resources to identify possible devices and establish their own standards for qualification testing (3). Furthermore, NEST360’s strong interdisciplinary expertise was key in identifying new devices for qualification testing, fostering collaboration with partners, such as UNICEF, and advocating for BMETs in facilities.

*Leadership and governance* barriers that continued despite NEST360’s intervention included exclusive distributor contracts, and import regulations, which led to shipping and import delays. Furthermore, there is currently no central regulatory authority for devices across the continent of Africa.

In *data systems and QI*, before NEST360 began to provide device support facilities were already tracking clinical outcomes so there was a precedent for tracking data. However, BMETs did not have their own system for monitoring data or indicators that were specific to their work. Additionally, QI projects historically were focused on clinical topics, and BMETs were not included in those projects.

Finally, prior to NEST360’s device implementation several barriers related to *infrastructure* prevented facilities from being able to consistently maintain functional devices, leading to equipment graveyards. This included harsh environmental conditions, such as power surges and the absence of heating, ventilation, and air conditioning systems, as well as simply not having enough space to store and perform maintenance on the devices. Some interviewees described how BMETs had to perform corrective or preventative maintenance under stairways or directly in the newborn unit since there was no other place for them to work in.

### 3.2 Strategies

Twenty-one strategies were implemented to address barriers to device identification, qualification, distribution, and management efforts. All identified strategies aligned with ERIC’s consolidated list (see Table 4). A detailed description of each strategy can be found in the supplemental material (Supplemental Table 1). Each strategy addressed multiple contextual factors, with strategies such as collaborating with stakeholders and developing Hatch Technologies targeting 17 factors and 11 factors respectively. Hatch Technologies is a device distribution and support company that facilitates the procurement of SSNC devices for facilities in sub-Saharan Africa and was developed by NEST360 after identifying the need for a device procurement and distribution system (28). More than half of the contextual factors (32 out of 40) were addressed by more than one strategy.

**Table 4.**
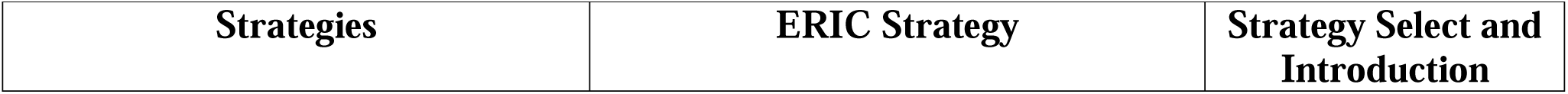
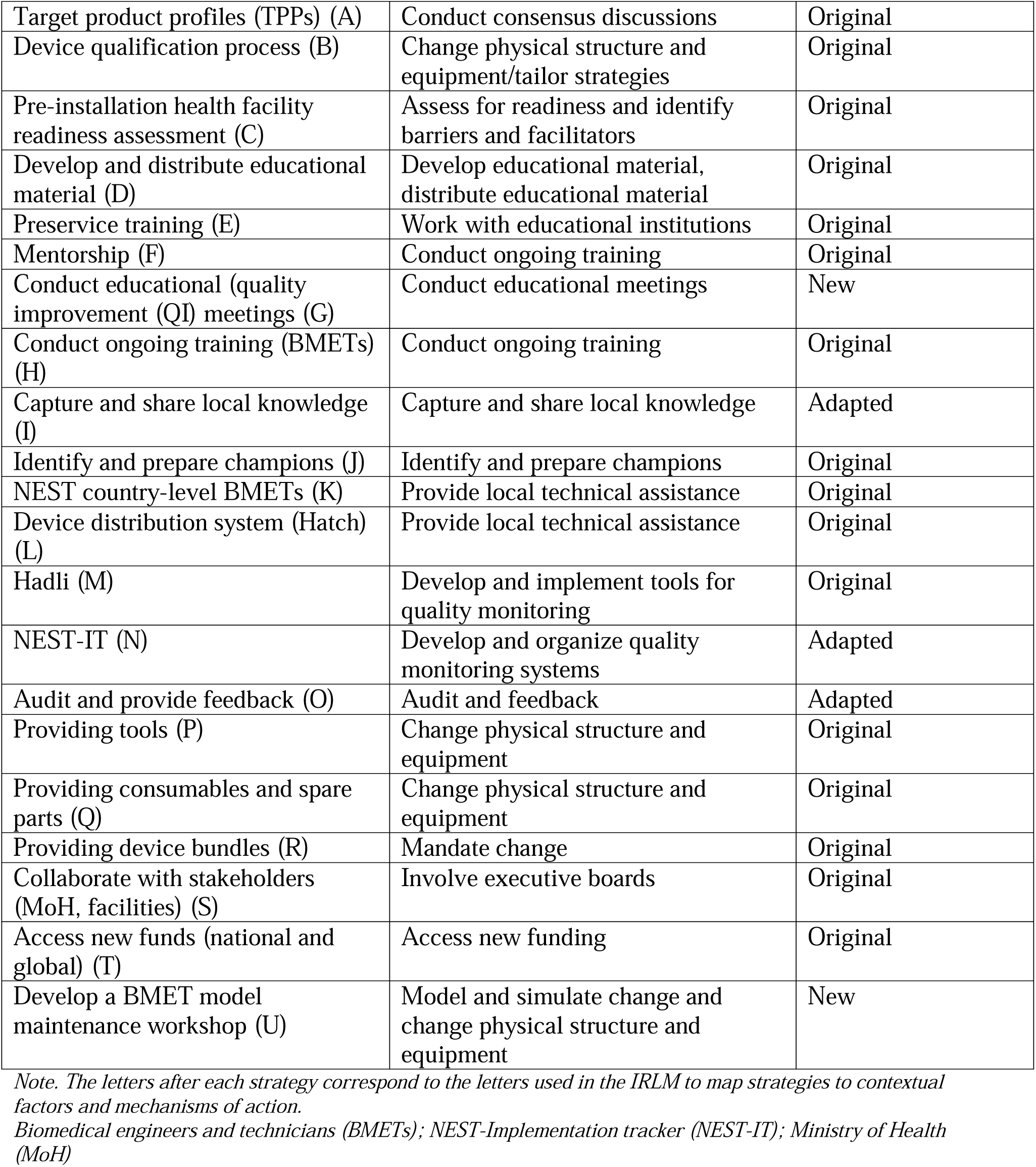
Inventory of strategies used to improve the implementation of device qualification, management, and distribution activities in NEST360 and their corresponding ERIC strategy.

| Strategies | ERIC Strategy | Strategy Select and Introduction |
| --- | --- | --- |
| Target product profiles (TPPs) (A) | Conduct consensus discussions | Original |
| Device qualification process (B) | Change physical structure and equipment/tailor strategies | Original |
| Pre-installation health facility readiness assessment (C) | Assess for readiness and identify barriers and facilitators | Original |
| Develop and distribute educational material (D) | Develop educational material, distribute educational material | Original |
| Preservice training (E) | Work with educational institutions | Original |
| Mentorship (F) | Conduct ongoing training | Original |
| Conduct educational (quality improvement (QI) meetings (G) | Conduct educational meetings | New |
| Conduct ongoing training (BMETs) (H) | Conduct ongoing training | Original |
| Capture and share local knowledge (I) | Capture and share local knowledge | Adapted |
| Identify and prepare champions (J) | Identify and prepare champions | Original |
| NEST country-level BMETs (K) | Provide local technical assistance | Original |
| Device distribution system (Hatch) (L) | Provide local technical assistance | Original |
| Hadli (M) | Develop and implement tools for quality monitoring | Original |
| NEST-IT (N) | Develop and organize quality monitoring systems | Adapted |
| Audit and provide feedback (O) | Audit and feedback | Adapted |
| Providing tools (P) | Change physical structure and equipment | Original |
| Providing consumables and spare parts (Q) | Change physical structure and equipment | Original |
| Providing device bundles (R) | Mandate change | Original |
| Collaborate with stakeholders (MoH, facilities) (S) | Involve executive boards | Original |
| Access new funds (national and global) (T) | Access new funding | Original |
| Develop a BMET model maintenance workshop (U) | Model and simulate change and change physical structure and equipment | New |

*Note. The letters after each strategy correspond to the letters used in the IRLM to map strategies to contextual factors and mechanisms of action.*
*Biomedical engineers and technicians (BMETs); NEST-Implementation tracker (NEST-IT); Ministry of Health (MoH)*

### 3.3 Outcomes

Results indicated a high level of implementation success (see Supplemental Table 2). NEST360’s device identification, qualification, and distribution efforts had a wide *reach*, with over 29 devices identified and qualified for twelve device categories for providing SSNC by the end of Phase 1 (2023). Further, 66 facilities received NEST-qualified devices and had them installed in their newborn units by the end of Phase 1 (2023). *Adoption* was also high with 2476 individual devices installed at NEST360-supported sites by the end of Phase 1 (2023).

NEST360’s implementation efforts were *effective*, as measured by the average number of newborn care devices that were functional at NEST360 facilities (both devices provided by NEST360 and other existing channels), and in 2024, 87% of devices were functional.

Additionally, 75% of all NEST360-supported facilities had enough functional CPAP devices to cover all eligible babies (Kenya: 69.2%, Malawi: 70.3%, Nigeria: 81.8%, Tanzania: 100%) by the end of Phase 1 (2023). Furthermore, 76.5% of all NEST360-supported facilities had enough functional phototherapy devices to cover all eligible babies by the end of Phase 1 (2023) (Kenya: 61.5%, Malawi: 81.1%, Nigeria: 90.1%, Tanzania: 57.1%). Interviewees reported that these high outcomes were influenced by in-person mentorship and telementorship from NEST360’s BMETs which helped facilities ensure device functionality and reduce downtime.

Through qualitative interviews with NEST360’s BMETs, we found high *acceptability* of NEST360’s device training for both clinicians and BMETs as well as positive reports related to the mentorship and support that NEST360 provided to facility employees. However, some interviewees reported uncertainty surrounding the steps taken within the device identification process as well as the timeline for qualifying devices. These interviews also identified several *adaptations* that the device identification, qualification, distribution, and management process underwent following the first five years of NEST360’s work. Some of these adaptations included opening a device qualification site in Malawi, and in some cases, transitioning BMET mentorship from in-person to telementorship.

*Feasibility* for maintaining functional devices at NEST360-supported facilities was high with 93% of these sites having a BMET available during the day or night shift to perform preventative or corrective maintenance by the end of Phase 1 (2023) (Kenya: 100%, Malawi: 86%, Nigeria: 100%, Tanzania: 100%). Interview participants reported that the pre-installation health facility assessment and data platforms helped to improve the feasibility of device installation and management. Furthermore, the NEST360 team demonstrated the feasibility of conducting device qualification testing outside the United States through their testing process in Malawi. Some barriers hindering feasibility remained related to exclusive distributor contracts and country-level import regulations, but overall NEST360, in partnership with Hatch, was able to import qualified devices into all NEST360-affiliates countries and install them at the facilities.

NEST360 had high *fidelity* related to the device qualification process because all 29 devices that were qualified underwent the same thorough process to receive their qualification approval. Additionally, the percentage of facilities performing monthly PPM for five of the six reported devices (glucometers, pulse ox, CPAP, oxygen concentrators, radiant warmers, and phototherapy) increased from 58.6% in 2022 (2/22-12/22, data from 1/22 was unavailable) to 70.8% in 2023 (1/23-12/23).

Our evaluation of the *maintenance* and s*ustainability* of NEST360’s device activities demonstrated continued improvement in PPM. From January to June 2024, the percentage of facilities conducting monthly PPM for five reported devices was 81.9%, demonstrating both stability and improvement in maintenance practices. Furthermore, interview participants reported that facility-level BMETs are independently performing both PPM and corrective maintenance. Participants also emphasize Hatch’s strong positioning for long-term sustainability beyond NEST360 activities, citing its expertise in identifying new technologies, robust data systems, quick partner response times, and its transition to a hybrid model integrating commercial sales with program support.

The qualitative interviews highlighted both a need for additional strategies to address the sustainment of program efforts as well as key actions NEST360 can take to address the issue of sustainability. These strategies included securing long-term funding for ACS, finalizing decommissioning policies, encouraging increased BMET staffing, ensuring NEST360-qualified devices and ACS are included on national device lists, and progressively transitioning responsibilities from NEST360 team members to facility staff.

### 3.4 Mechanisms of action and the implementation research logic model

Using the IRLM (Figure 2), we mapped the relationship between contextual factors, strategies, and implementation outcomes of NEST360’s device qualification, management, and distribution work. For example, through the development of target product profiles (TPPs) and the device qualification process, NEST360 was able to ensure that the devices used at the facility level were affordable, accessible, and tailored for use in resource-limited settings, which enables the feasibility and sustainability of functional devices at these facilities. Additionally, through the creation of Hatch Technologies, NEST360 was able to support distributors with getting their products into NEST360’s partner countries which increased the feasibility of getting devices to the facilities and maintaining functionality through ongoing service. This also supports long-term availability of devices as Hatch continues as an independent distribution organization that facilities can continue to procure devices from post NEST360 involvement. All mechanisms of action for this work are presented in Figure 2.

**Figure 2.**
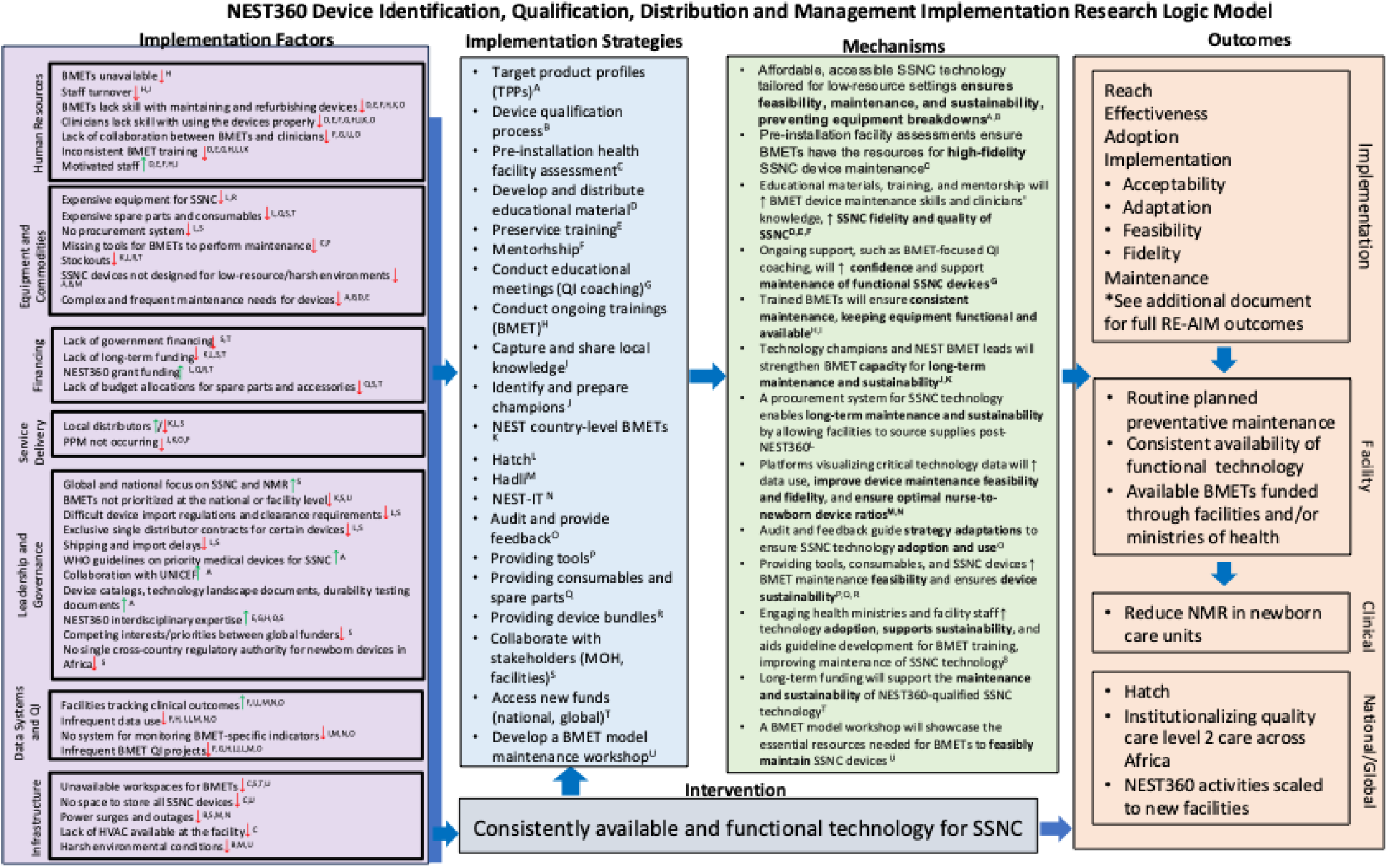
**An implementation research logic model for device qualification, distribution, and management activities within NEST360**. *Note: Biomedical engineers and technicians (BMETs); Small and sick newborn care (SSNC), Planned preventative maintenance (PPM); Neonatal mortality ratio (NMR); World Health Organization (WHO); United Nations International Children’s Emergency Fund (UNICEF); Quality improvement (QI); Health, ventilation, and air conditioning (HVAC); NEST-Implementation Tracker (NEST-IT); Ministry of health (MOH)*

## 3. Discussion

This study explored NEST360’s device identification, qualification, distribution and management activities through an implementation research lens. We identified 40 contextual factors, which mapped to 21 implementation strategies, and found strong evidence of positive emerging implementation outcomes. Through the identified strategies, NEST360 has been able to ensure facilities have access to functional, affordable medical devices while working on long-term sustainability through partnerships, mentorship, and innovative comprehensive distribution models through Hatch Technologies.

NEST360’s strategies effectively addressed multiple challenges related to implementing and maintaining medical devices by involving BMETs in QI initiatives, equipping them with progress indicators, and ensuring device sustainability for local environments. Simple yet impactful solutions, such as providing toolboxes were essential for effective maintenance.

However, sustaining progress requires ongoing training and mentorship to build long-term capacity, as well as financial sustainability through government buy-in, long-term funding mechanisms, and proactive budgeting before NEST360 funding phases out. Addressing distribution barriers as well as policy reform and advocacy will be key to improving the availability of functional devices and institutionalizing NEST360’s efforts

This study provides an implementation-focused analysis of strategies to ensure functional devices for SSNC in resource-limited settings. Tools like the IRLM, RE-AIM, and SSNC implementation toolkit help identify barriers, facilitators, and strategies. It also demonstrates how IR methods can be integrated into program evaluation to drive change and prioritize actions (29).

Given the gap between clinical innovation and sustainable implementation (30), the study highlights mechanisms like Hatch Technologies and NEST360’s mentorship model as scalable solutions for transitioning from donor-driven to locally managed systems. By combining qualitative and quantitative methods, it offers an evidence-based roadmap for overcoming systemic challenges in device implementation with globally relevant solutions.

This work identified simple yet effective strategies for enhancing neonatal care and device maintenance. Collaboration between BMETs and clinicians to address functionality challenges and align QI priorities was seen across all four NEST360 countries, and aligns with previous research highlighting the role of BMETs and importance of multidisciplinary teams’ in enhancing clinical care and preventative maintenance (10, 31). Additional strategies that align with the current literature, include hands-on training for BMETs and clinicians with devices like CPAP and phototherapy machines to build technical expertise and ensure proper use, and leveraging tools like the NEST-IT dashboard for monitoring device performance, guiding resource allocation, and informing maintenance schedules (32, 33). Finally, dedicated storage and maintenance spaces should be established to enhance organization and efficiency (34). Implementing these strategies can improve neonatal device functionality, strengthen care systems, and enhance clinical outcomes for newborns.

While 21 implementation strategies were employed by NEST360, the full package may not be required for other programs and settings. It is likely that future programs may not require the full set of strategies to achieve similar results, particularly if certain enabling factors-such as preservice training systems or pre-developed tools like TPPs are already in place. Tailoring strategy selection to local context and available infrastructure can enhance efficiency without compromising implementation quality.

Participant policy recommendations emphasize the critical need to strengthen human resources by prioritizing the recruitment, training, and retention of BMETs and including them in national health workforce planning. Sustainable funding for ACS, and maintenance should be secured through facility budgets and innovative financing models. Both recommendations align with WHO recommendations and the current literature, which underscore the importance of maintaining trained BMETs and ensuring the sustainability of ACS to ensure the functionality of devices (35–38). Additionally, an Africa-wide regulatory authorities should be developed to oversee level 2 SSNC medical devices to ensure they meet quality, durability, and safety standards suitable for resource-limited settings. This would be similar to the recent decision made by Africa’s World Health Organization Maturity Level 3 National Regulatory Authorities who signed a Memorandum of Understanding (MoU) to enhance collaboration and expedite the approval processes for medicines, vaccines, and medical devices across the continent. A similar alignment of regulations regionally may help with the challenges identified with importation of level 2 SSNC devices (39). Addressing these policy areas will strengthen healthcare systems, support neonatal care, ensure device sustainability, and achieve long-term reductions in neonatal mortality.

These findings highlight critical research implications, particularly for improving neonatal care and medical device maintenance in low-resource settings. Future studies will explore adapting and scaling NEST360’s strategies for device functionality at additional sites and assessing the long-term sustainability of initiatives like mentorship program and the Hatch model post-external support. Additionally, this study demonstrated how IRLMs can be used in resource-limited settings to articulate the relationship between implementation strategies and outcomes, which aligns with previous studies that have benefitted from mapping their global projects and programs to an IRLM (40–42). Finally, the IRLM can be used as a guide for NEST360’s Phase 2 (2024-2029) activities. The IRLM is useful for identifying key barriers and facilitators that influenced implementation success in Phase 1 and linking strategies to mechanisms to understand what drove that change and where gaps remain. Furthermore, this tool can help identify what to monitor next, including which determinants to revisit or track in phase 2 and prioritize strategies that were most impactful or adaptable. Future research should address inconsistent PPM performance and strategies for improving adherence. Additionally, innovative financing models for device maintenance and comparative studies on the broader applicability of NEST360’s approaches beyond neonatal care are needed.

## Limitations

This study was unable to establish a direct correlation between specific implementation strategies and improved newborn outcomes, as the clinical impact of NEST’s device activities cannot be isolated from their additional education, quality improvement, and capacity building activities. Furthermore, this is a short-term evaluation, which does not comprehensively assess long-term sustainability. However, we will continue to use IR outcomes to explore NEST360’s sustainability as well as measure implementation success during scale up to new level 2+ neonatal units. Additionally, our analysis did not include grading the relative influence of contextual barriers and facilitators (e.g., assigning weighted values such as -2 to +2), which may have provided additional insight for prioritizing implementation strategies; future work will explore this approach to better tailor interventions to the magnitude of influence. Finally, while the core functions of implementation strategies were consistent across countries, there were variations in form, dose, and temporality due to contextual factors (e.g., health worker strikes, natural disasters); detailed analysis of these adaptations is the focus of ongoing work.

## 4. Conclusions

In conclusion, this study highlights how NEST360 successfully implemented and maintained functional medical devices in resource-limited settings by addressing key contextual factors through targeted strategies. The application of IR in this article allowed for a deeper understanding of how these strategies aligned with the local context, providing insights into areas requiring improvement to ensure sustained device access and functionality. The findings and methods from this work will inform the scale-up of NEST360 device activities and can be used as a blueprint by other programs for sustaining device availability and functionality in resource-constrained healthcare systems globally.

## Data Availability

The dataset used during the current study is fully owned by the facilities which provided the information. Data that could be linked to a particular facility will not be shared outside of NEST. Aggregated data (multi-country and program level) may be made available at the discretion of the NEST360 Steering Committee. Requests for data access can be made on the NEST360 website in the Request for Public Use of NEST360 Data section. Many of the tools and educational materials mentioned in this manuscript are already available on the NEST360 website in the Resources section. Requests for additional tools, material, and learnings are available from the NEST360 Steering Committee upon reasonable request.

https://nest360.org/data-use/

## List of abbreviations

SSNC: Small and sick newborn care
CPAP-: Continuous positive airway pressure
NEST360: Newborn Essential Solutions and Technologies
BMETs: Biomedical engineers and technicians
QI: Quality improvement
IRLM: Implementation research logic model
UNICEF: United Nations Children’s Fund
ERIC: Expert Recommendations for Implementation Research
NEST-IT-NEST: Implementation Tracker
PPM: Planned preventative maintenance
KII: Key informant interviews
ACS: Accessories, consumables, and spare parts
TPPs: Target product profiles

## 6. Declarations

### 7.1 Ethics approval and consent to participate

Ethics approvals were obtained from the London School of Hygiene and Tropical Medicine IRB (21892), and the national health research ethics committees in Malawi (NHSRC 1180), Kenya (Kenya Medical Research Institute: KEMI/SERU/CGMR-C/229/4203, Maseno University Ethics Review Committee: MSU/DRPI/MUERC/00810/19), Tanzania (Ifakara Health Institute: IHI/IRB/No:01-2020, National Institute for Medical Research: NIMR/HQ/R.8a/Vol.IX/3405), and Nigeria (Lagos University Teaching Hospital Health Research Ethics Committee: ADM/DCST/HREC/APP/3487, University of Ibadan/University College Hospital/Ethics Committee: NHREC/05/01/2008a). This work used pre-existing programmatic data that was de-identified. This specific study was deemed program evaluation work and not human subjects research and thus Northwestern waived the requirement of an IRB review.

### 7.2 Consent for publication

Not applicable as no individual-level data are presented.

### 7.3 Availability of data and materials

The dataset used during the current study is fully owned by the facilities which provided the information. Data that could be linked to a particular facility will not be shared outside of NEST. Aggregated data (multi-country and program level) may be made available at the discretion of the NEST360 Steering Committee. Requests for data access can be made on the NEST360 website in the <u>Request for Public Use of NEST360 Data section</u>. Many of the tools and educational materials mentioned in this manuscript are already available on the NEST360 website in the <u>Resources section</u>. Requests for additional tools, material, and learnings are available from the NEST360 Steering Committee upon reasonable request.

## 7.6 Authors’ contributions

This work was done in partnership with the NEST360 Alliance. The author’s views are their own, and not necessarily from any of the institutions they represent. KP, RK, EA, MA, DK, VG, NB, AI, RRK, ZMO, and CB made substantial contributions to the conception of this work. KD, KC, KP, RK, CB, and LRH made significant contributions to the design of this work. KC cleaned all transcripts and analyzed the data with KD. KD, KC, KP, RK, EA, MA, DK, VG, NB, AK, VOO, GB, HDP, CO, RRK, ZMO, CB, and LRH made substantial contributions to the acquisition and interpretation of the data. KD drafted the work with substantial revisions from RK, and LRH. All authors read and approved the final manuscript and agree to be personally accountable for the author’s own contributions and to ensure that questions related to the accuracy or integrity of any part of the work, even ones in which the author was not personally involved, are appropriately investigated, resolved, and the resolution documented in the literature.

## 7.7 Acknowledgements

First, and most importantly, we thank the newborns and their mothers whose data are the heart of NEST360. We also thank the biomedical technicians and clinicians involved in the day-to-day device management and utilization activities. Many thanks to the relevant administrative staff for their support. We are also very grateful to the fellow researchers and editors who peer-reviewed this paper and for the input from the managing editors at *BMC Global and Public Health* and within NEST360.

---

**Supplemental Table 1.**
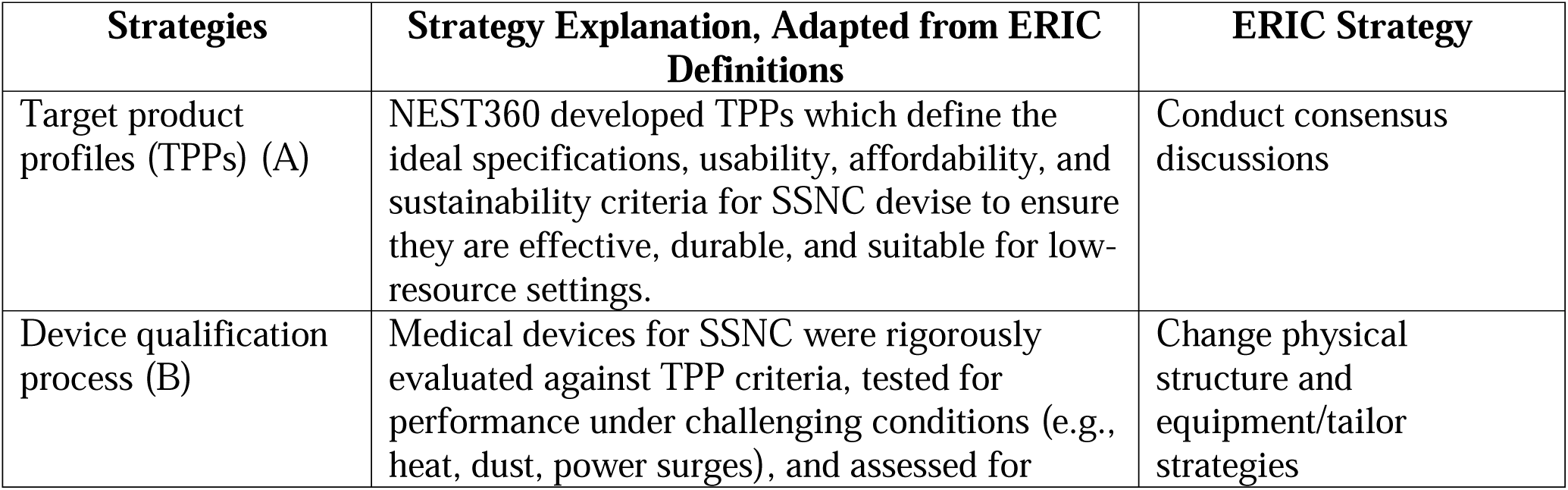
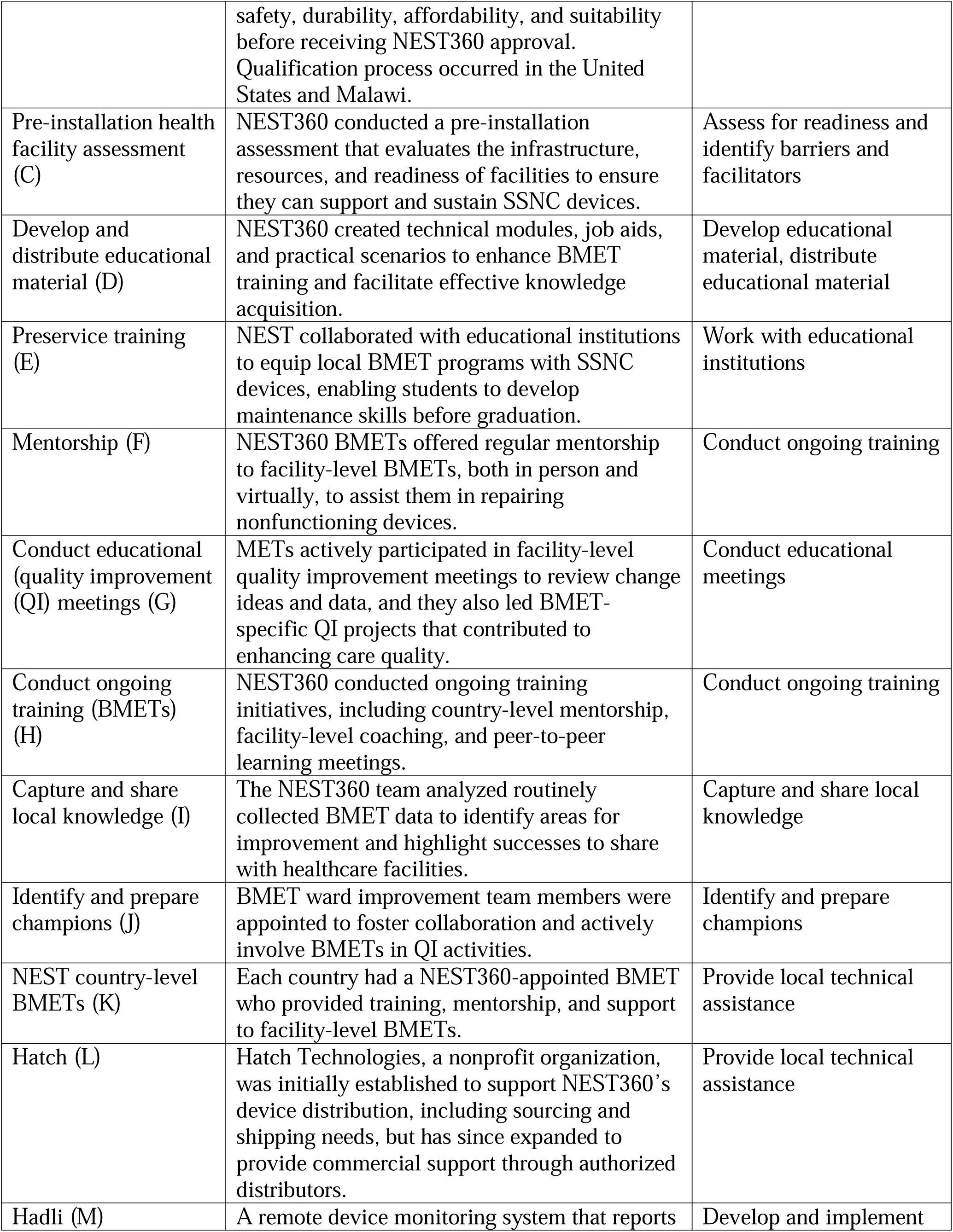
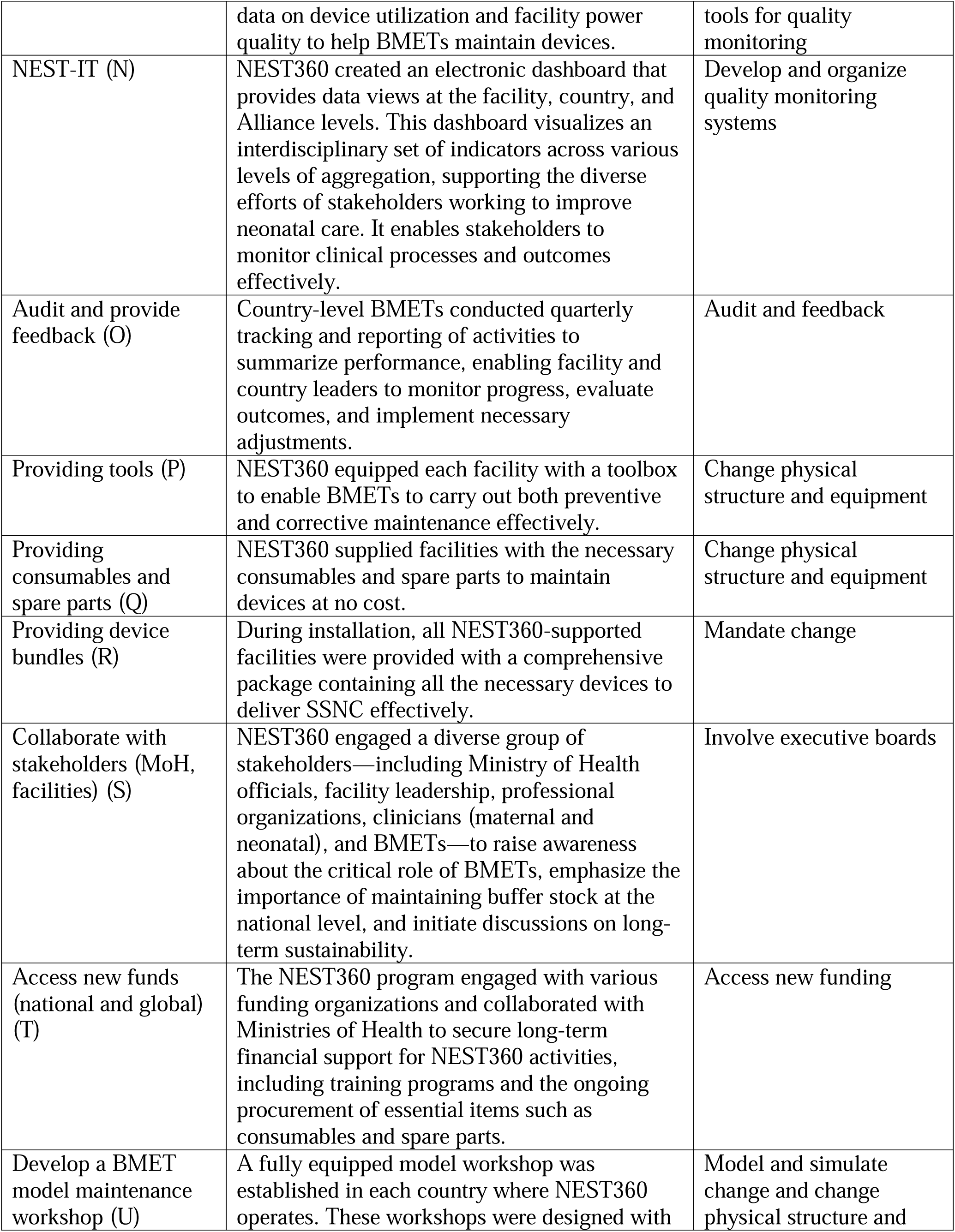
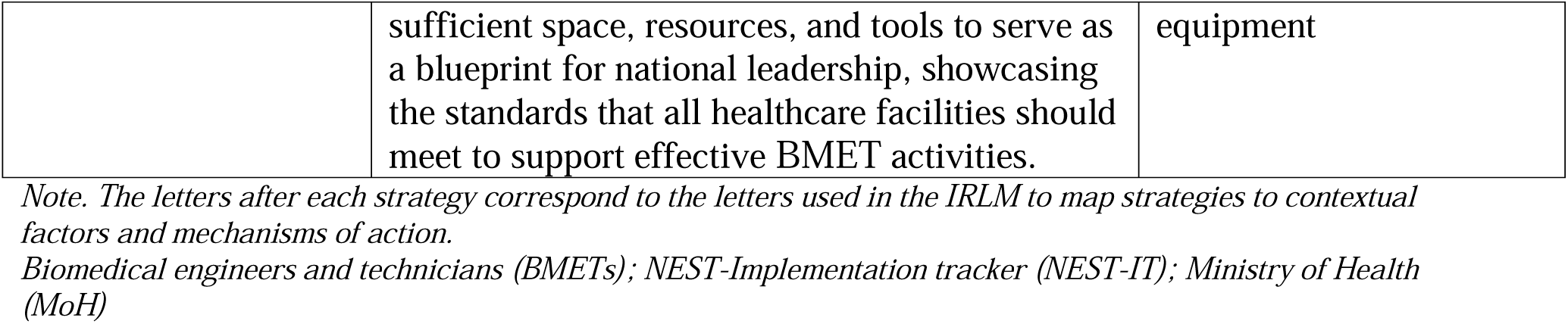
Inventory of strategies used to improve the implementation of device qualification, management, and distribution activities in NEST360.

**Supplemental Table 2.**
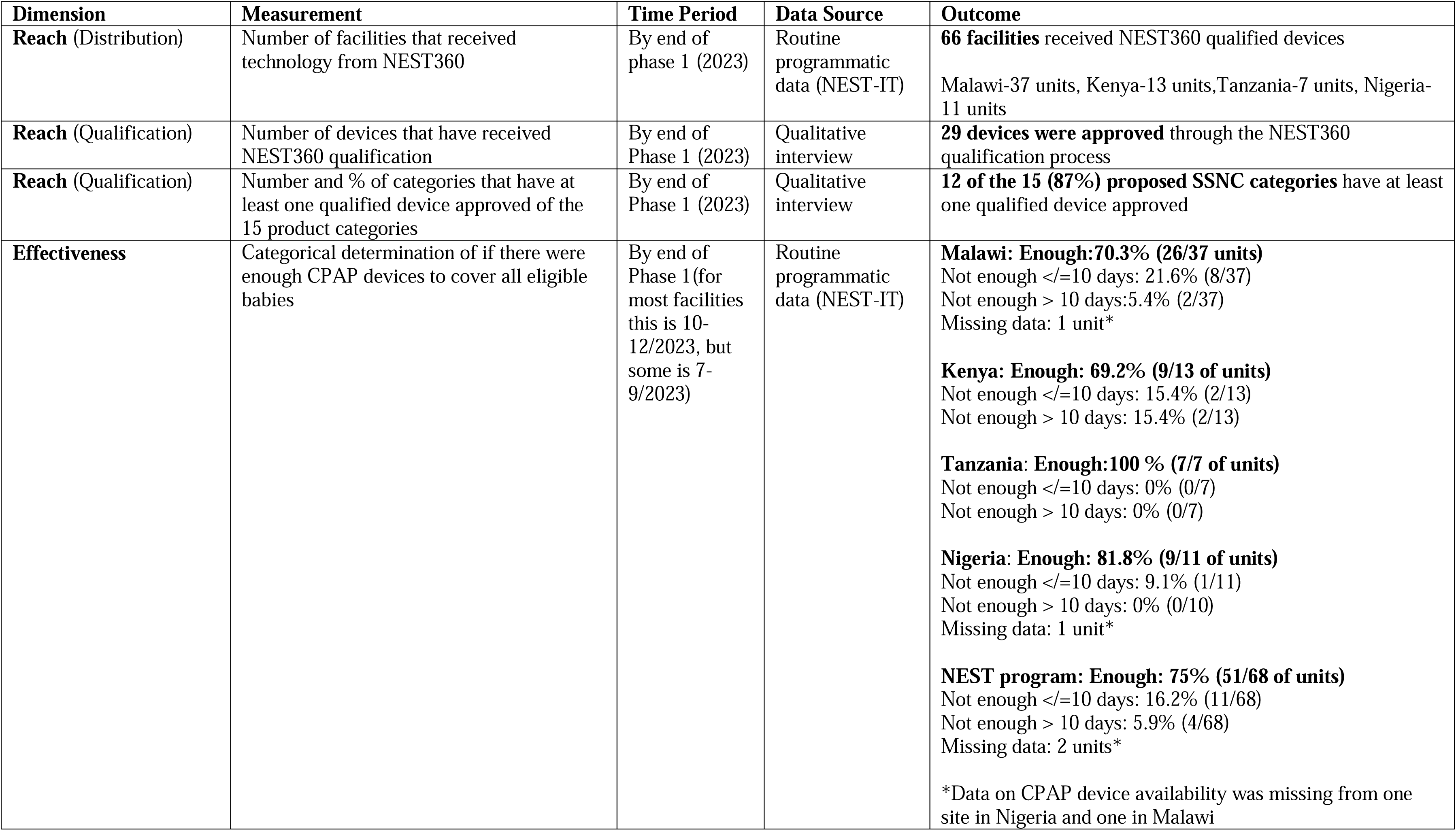
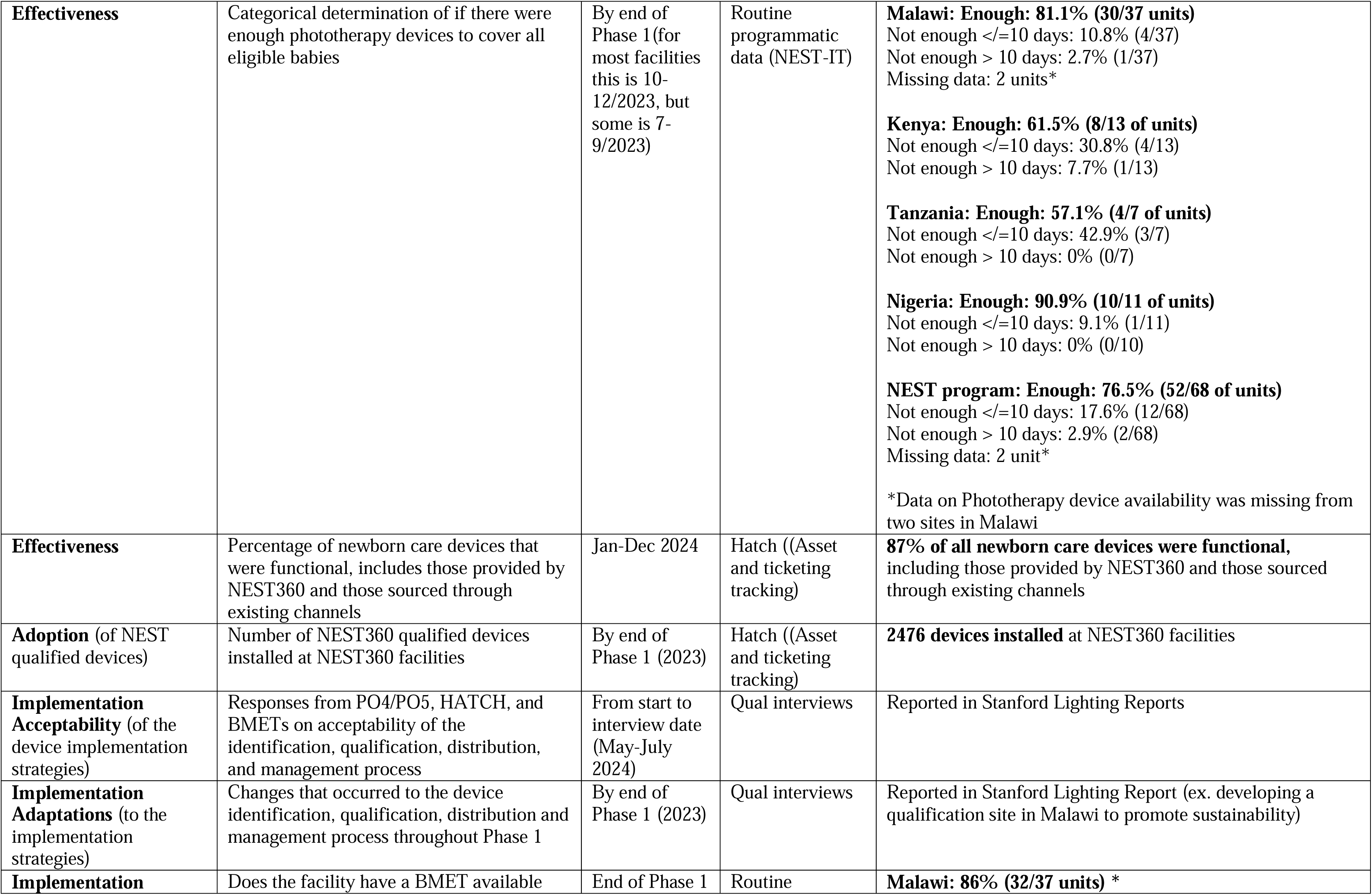
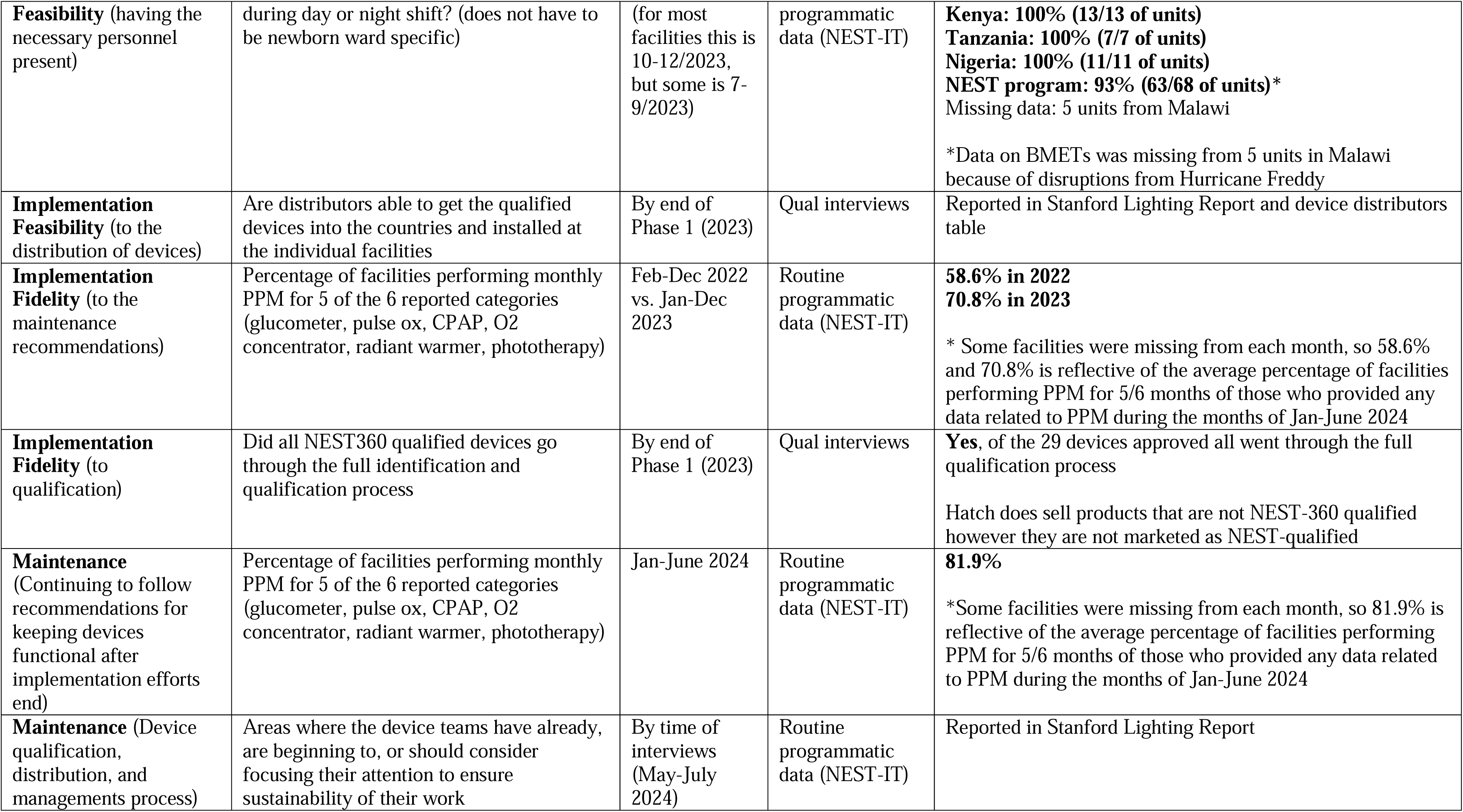
Detailed Definitions of NEST360’s Device Identification, Qualification, Management, and Distribution Implementation Outcomes.

