## Supplemental material 2 for "From warehouse to ward: Applying implementation research methods to the device identification, qualification, distribution, and management process within the NEST360 alliance"

**Qualitative Interview Guide for PO4 Technology Implementation**

The purpose of this interview is to explore and describe the selection and implementation of strategies for qualifying technology and implementing it at the facility level. For this work, the focus is on the work related to the identification, implementation, and maintenance of appropriate technology needed to implement evidence-based interventions (EBI) known to reduce neonatal mortality in small and sick newborns in level 2 nurseries in NEST360.

We are interested in hearing your opinions and learning about your personal experiences. There are no right or wrong answers, we are interested in your honest opinion, and everything you say during this interview/discussion will be kept confidential.

Ask permission for an audio recording of the interview for further analysis and record interview accordingly. All audio files will be stored on a password protected OneDrive and will be destroyed following transcription. All transcripts will be deidentified prior to analysis.

First, I want to ask you a few questions about yourself.

1. What is your job title?
2. What is your role?
3. Years of experience with this position? (more specific for BMETs and distributors)

Device qualification team specific questions

1. Can you describe how you developed the process?
   a. Newborn technology landscape document
   b. Lots of googling looking for new products, USAID reviewed products, WHO technology compendium and now leveraging HATCH knowledge, VIA global health international database
2. Are you constantly looking for new documents or only when issues are identified with current products?
3. Do national policies play a role in the qualification process?
4. Can you describe the factors influencing the qualification process?
   a. Are there common barriers to identifying and qualifying devices?
   b. Are there lots of equipment options available/only a few/none?
5. Does Rice plan to test forever or hopes to transition technical testing to Africa?
6. How did you determine the environmental conditions?
   a. How did you determine level of heat/dust?
   b. Military durability standards
   c. TPP environmental standards
7. Any barriers with conducting usability testing in Houston and/or Malawi?
8. Can you describe the thought process that went into the decisions to focus on [respiratory products] category for SSNC?
   a. Was it based on country-led or facility-led requests or based on WHO recommendations/guidelines?
   b. Why were respiratory products viewed as a higher priority than other medical devices?
   c. Any policies or regulations influencing this?
9. Is there anything else that you’d like me to know about the topics that we’ve discussed today that I didn’t ask about?

**Qualitative Interview Guide for PO5 Technology Implementation**

I would like to ask you a few general technology implementation questions.

1. Can you describe the process for how technology was introduced to the facilities?
   a. Signing agreements with suppliers?
   a. What were the barriers and facilitators to the implementation process?
   b. What strategies were used? How were they selected and adapted?
   c. Which individuals are involved in the implementation process?
   d. What was the training like?
   e. How does this differ based on kind of equipment?
   f. What other NEST360 teams do you work with?
   g. How and when were facility staff involved?
2. What factors prevent devices from being used right away?
3. How are staff supported at the facilities to maintain technology?
4. What information do you collect to track BMET issues and strengths?
5. What do you see as the major challenges to maintaining functional equipment at the NEST360 sites?
   a. In the short term
   b. In the long term
6. How do you measure successful implementation of equipment?
   a. What factors influenced these outcomes?
   b. How does this differ by type?
7. What do you see as the major challenges to have the right equipment available?
   a. What about functional at facilities?
   b. Ex. Procurement, maintenance, facility resources (power outages)
8. Transitioning activities for BMETs
9. Is there anything else that you’d like me to know about the topics that we’ve discussed today that I didn’t ask about?

**Qualitative Interview Guide for BMETs on Technology Implementation**

Now I would like to ask you a few general technology implementation questions.

1. Are you involved in the procurement and introduction of devices to the facility?
2. Can you describe the process for how technology was introduced to the facilities?
   h. What were the barriers and facilitators to the implementation process?
   i. What strategies were used? How were they selected and adapted?
   j. Which individuals are involved in the implementation process?
   k. What was the training like?
   l. How does this differ based on kind of equipment?
   m. How and when were facility staff involved?
3. How are staff supported at the facilities to maintain technology?
   a. Leadership, frequent reminders, routine rounds on the neonatal wards?
4. What do you see as the major challenges to maintaining functional equipment at the NEST360 sites?
   a. In the short term
   b. In the long term
5. What do you see as the major challenges to have the right equipment available and functional?
   a. Maintenance
   b. Not enough devices
   c. Not enough BMETs/nurses
   d. Cost of spare parts/replacements
   e. Facility resources (power outages), procurement, maintenance
6. Do you feel the facilities under your jurisdiction have adequate space to store and use the equipment?
   a. Is there space for transfer as well?
7. Can you describe steps currently being taken to support BMET sustainability following the conclusion of NEST360 phase 2?

**Qualitative Interview Guide for Hatch on Technology Implementation**

1. How do you identify the devices that you want to be qualified?
2. Can you tell me about how you market NESt products?
3. What is the process for retiring a device?
   a. Is it just calling it decommissioned or do you go and remove the devices?
   b. How are they disposed?
4. Can you describe how Hatch supports facilities with the process of ordering and receiving supplies?
   a. Barriers and facilitators?
   b. How do you handle the intra Africa distribution?
   c. Things you like/dislike?
   d. Country specific factors?
   e. Program specific factors?
5. What are the benefits of working with Hatch?
6. Are there different financial reporting for each NEST country?
7. Are there specific factors that impact distribution of respiratory devices?
   a. Higher costs, more regulations on import, more fragile parts?
8. How do you get info and details related to device functionality from the sites?
   a. If you aren’t, how would you like to?
9. What factors about HATCH support their sustainability after phase 2?
10. Of all the NEST360 qualified technology, which items are difficult/not possible for you to procure?
11. Is there anything else that you’d like me to know about the topics that we’ve discussed today that I didn’t ask about?
