## Supplemental material 3 for "From warehouse to ward: Applying implementation research methods to the device identification, qualification, distribution, and management process within the NEST360 alliance"

| **Code/Domain** | **Definition** | **Subcodes** | **When to apply** | **When not to apply** |
| --- | --- | --- | --- | --- |
| Human resources | The dedicated workforce needed to provide neonatal care including doctors, neonatal nurses, biomedical technicians/engineers (BMETs), effective hospital management, and other support staff in the proper ratios. These teams require the right mix of interdisciplinary skills, competencies and knowledge to support the needs of small and sick newborns and their families. | Facilitators | - When participants describe the people and teams that make it easier to provide small and sick newborn care (SSNC) - When participants describe the educational needs and resources including pre-service and in-service trainings that support their ability to provide SSNC - When participants describe the enabling environment, including mentorship and supportive supervision to ensure the workforce is motivated, adequately compensated and able to implement quality improvement (QI) approaches | - When participants describe components outside of the workforce dedicated to supporting SSNC, their educational and skill needs and resources or their enabling environment - When participants describe factors related to the workforce, educational needs and resources that make it harder to provide SSNC |
| Human resources | The dedicated workforce needed to provide neonatal care including doctors, neonatal nurses, biomedical technicians/engineers (BMETs), effective hospital management, and other support staff in the proper ratios. These teams require the right mix of interdisciplinary skills, competencies and knowledge to support the needs of small and sick newborns and their families. | Barriers | - When participants describe the people and teams that make it harder to provide SSNC - When participants describe the educational needs and resources including pre-service and in-service trainings that are not available but would be necessary to provide high quality SSNC - When participants describe environmental factors, such as supportive supervision or mentorship that are not available or lacking which led them to not be adequately motivated, compensated or able to implement QI approaches | - When participants describe components outside of the workforce dedicated to supporting SSNC, their educational and skill needs and resources or their enabling environment - When participants describe factors related to the workforce, educational needs and resources that make it easier to provide SSNC |
| Leadership & governance | Effective leadership and governance require a participatory approach through community empowerment and accountability, along with engagement with the right stakeholders such as key-decision makers, policy influencers, funders, clinicians, and hospital management, from various disciplines at the national, district and hospital levels. | Facilitators | - When participants describe engagement of stakeholders for effective prioritization of newborn health, identifying national targets, and creating long-term policy frameworks to support SSNC - When participants describe effective participatory leadership through empowerment of service users and members of the community - When participants describe methods for ensuring quality care by communicating of goals and priorities through policies and guidelines, and gathering information to track whether goals are being met - When participants describe methods for ensuring financial, performance, or political accountability | - When participants describe components outside of stakeholder engagement, participatory leadership, guidelines, quality of care governance, or accountability - When participants describe factors related to the stakeholder engagement, participatory leadership, guidelines, quality of care governance, or accountability that make it harder to provide SSNC |
| Leadership and governance | Effective leadership and governance require a participatory approach through community empowerment and accountability, along with engagement with the right stakeholders from various disciplines at the national, district and hospital levels. Stakeholders include key decision-makers, policy influencers, funders, clinicians, and hospital management. | Barriers | - When participants describe ineffective or nonexistent engagement of stakeholders that does not help to promote prioritization of newborn health, identifying national targets, and creating long-term policy frameworks to support SSNC - When participants describe a lack of participatory leadership through empowerment of service users and members of the community - When participants describe ineffective or a lack of methods for ensuring quality care by communicating of goals and priorities through policies and guidelines, and gathering information to track whether goals are being met - When participants describe ineffective methods for ensuring financial, performance, or political accountability | - When participants describe components outside of stakeholder engagement, participatory leadership, guidelines, quality of care governance, or accountability - When participants describe factors related to the stakeholder engagement, participatory leadership, guidelines, quality of care governance, or accountability that make it easier to provide SSNC |
| Data systems & Quality improvement (QI) | Timely, accurate data from standardized monitoring is needed to highlight success and identify where the focus is needed | Facilitators | - When participants describe an effective Routine Health Information systems (RHIS): any data collection, analysis, and dissemination occurring at regular intervals (of <1 year) to meet predictable information needs - When participants describe easy availability of information regarding the content and quality of care at all health system levels - When participants describe the use of any global indicator data system such as The Every Newborn Action Plan, World Health Organization (WHO) 100 Core Health Indicators, and (MoNITOR) - When participants describe evidence-based decision making or use of QI initiatives to inform practice, organization, and delivery of care. | - When participants describe components outside of a Routine Health Information system, availability of information, global indicator systems, or QI initiatives - When participants describe factors related to the Routine Health Information system, availability of information, data systems, or QI initiatives that make it harder to provide SSNC |
| Data systems & QI | Timely, accurate data from standardized monitoring is needed to highlight success and identify where the focus is needed. | Barriers | - When participants describe an ineffective or missing RHIS - When participants describe challenges to obtaining information regarding the content and quality of care at all health system levels - When participants describe factors that prevent them from making evidence-based decision, or using QI initiatives to inform practice, organization, and delivery of care. | - When participants describe components outside of a Routine Health Information system, availability of information, data systems, or QI initiatives - When participants describe factors related to the Routine Health Information system, availability of information, global indicator systems, or QI initiatives that make it easier to provide SSNC |
| Referral system | A well-functioning referral system links early detection of severe illness in newborns with the subsequent appropriate advanced care so that newborns and their mothers can receive care of the highest quality and at the lowest cost at the most appropriate level. | Facilitators | - When participants describe referral strategies or protocols which effectively consider population needs and health system capabilities and make it easier to provide SSNC - When participants describe sufficient infrastructure pertinent to the referral system including a referral center, designated transport, trained personnel at each personnel level, and a unified records system - When participants describe effective collaboration between referral levels and across sectors - When participants describe a strategy to ensure continuity within record systems and accountability for provider performance | - When participants describe components outside of a referral strategy, referral protocols, referral infrastructure, collaboration between referral levels, and medical record systems - When participants describe infrastructure components that influence of the delivery of SSNC but do not pertain to the referral system - When participants describe factors related to a referral strategy, referral protocols, referral infrastructure, collaboration between referral levels, and medical record systems that make it harder to provide SSNC |
| Referral system | A well-functioning referral system links early detection of severe illness in newborns with the subsequent appropriate advanced care so that newborns and their mothers are able to receive care of the highest quality and at the lowest cost at the most appropriate level. | Barriers | - When participants describe an ineffective referral strategy which does not meet population needs or account for health system capabilities - When participants describe a lack of infrastructure pertinent to the referral system including a referral center, designated transport, trained personnel at each personnel level, and a unified records system - When participants describe ineffective protocols of referral systems that make it harder to provide SSNC - When participants describe ineffective collaboration between referral levels and across sectors - When participants describe the lack of accountability for provider performance | - When participants describe components outside of a referral strategy, referral protocols, referral infrastructure, collaboration between referral levels, and medical record systems - When participants describe infrastructure components that influence of the delivery of SSNC but do not pertain to the referral system - When participants describe factors related to a referral strategy, referral protocols, referral infrastructure, collaboration between referral levels, and medical record systems that make it easier to provide SSNC |
| Infrastructure | Infrastructure needed to support quality small and sick newborn care includes adequate physical space, water, sanitation, and hygiene (WASH) facilities, reliable electrical power, medical gases and vacuum systems, and referral and transportation systems. | Facilitators | - When participants describe sufficient physical space to support staff and families, along with practices of Kangaroo mother care, breastfeeding, and infection control - When participants describe appropriate water supply, hand hygiene stations, toilets, washing facilities, disinfection protocols, waste management, and heating, ventilation, and air conditioning - When participants describe adequate access to reliable, high quality electrical power - When participants describe adequate infrastructure to support medical oxygen, medical air, and suction systems to support medical devices - When participants describe adequate infrastructure to support referral system levels and transportation from home to the hospital, from primary health centers to the hospital and between hospitals | - When participants describe components outside of infrastructure for adequate physical space, water, sanitation, and hygiene (WASH) facilities, reliable electrical power, medical gases and vacuum systems, and referral and transportation systems - When participants describe factors related infrastructure for adequate physical space, water, sanitation, and hygiene (WASH) facilities, reliable electrical power, medical gases and vacuum systems, and referral and transportation systems that make it harder to provide SSNC |
| Infrastructure | Infrastructure needed to support quality small and sick newborn care includes adequate physical space, water, sanitation, and hygiene (WASH) facilities, reliable electrical power, medical gases and vacuum systems, and referral and transportation systems. | Barriers | - When participants describe insufficient physical space to support staff and families, along with practices of Kangaroo mother care, breastfeeding, and infection control - When participants a lack of appropriate water supply, hand hygiene stations, toilets, washing facilities, disinfection protocols, waste management, and heating, ventilation, or air conditioning - When participants describe inadequate access to reliable, high quality electrical power - When participants describe inadequate infrastructure to support medical oxygen, medical air, and suction systems to support medical devices - When participants describe inadequate infrastructure to support referral system levels and transportation from home to the hospital, from primary health centers to the hospital and between hospitals | - When participants describe components outside of infrastructure for adequate physical space, water, sanitation, and hygiene (WASH) facilities, reliable electrical power, medical gases and vacuum systems, and referral and transportation systems - When participants describe factors related infrastructure for adequate physical space, water, sanitation, and hygiene (WASH) facilities, reliable electrical power, medical gases and vacuum systems, and referral and transportation systems that make it easier to provide SSNC |
| Equipment & commodities | Quality care for small and sick newborns requires that essential medical supplies required for level 2 SSNC be readily available, dosed appropriately, at an affordable price with assured quality. The selection of medical supplies must consider both the needs and capacities of the local hospitals. | Facilitators | - When participants describe all the medications, supplies, and items (ex. KMC wraps) that are necessary to provide high quality SSNC - When participants describe the selection of medical supplies in the right quantities to meet the needs of the target population - When participants describe the presence of a strong supply chain for the proper usage and maintenance of medical devices - When participants describe an adequate system of assessment of medical technology needs and innovation for those needs. | - When participants describe components outside of the process of selection of appropriate medical technologies considering population needs, physical demand, input from and training of hospital staff, supply chain requirements, and innovation around such technologies - When participants describe factors related to the process of selection of appropriate medical technologies considering population needs, physical demand, input and training of hospital staff, supply chain requirements, or innovation around such technologies that make it harder to provide SSNC |
| Equipment & commodities | Quality care for small and sick newborns requires that essential medical supplies required for level 2 SSNC be readily available, dosed appropriately, at an affordable price with assured quality. The selection of medical supplies must consider both the needs and capacities of the local hospitals. | Barriers | - When participants describe factors that prevent them from having all the medications, supplies, and items (ex. KMC wraps) that are necessary to provide high quality SSNC - When participants describe the ineffective selection of medical supplies that do not meet the needs of the target population - When participants describe the lack of a strong supply chain for the proper usage and maintenance of medical devices - When participants describe an inadequate system of assessment of medical technology needs and innovation for those needs. | - When participants describe components outside of the process of selection of appropriate medical technologies considering population needs, physical demand, input and training of hospital staff, supply chain requirements, and innovation around such technologies - When participants describe factors related to the process of selection of appropriate medical technologies considering population needs, physical demand, input and training of hospital staff, supply chain requirements, or innovation around such technologies that make it easier to provide SSNC |
| Infection prevention & control | Infection Prevention and Control (IPC) practices, including the assurance of clean birth and care settings, early detection of neonatal infections, and appropriate treatment of neonatal infections in settings providing newborn care services, are critical for reducing neonatal infections. | Facilitators | - When participants describe training for and monitoring of best practices for the prevention of healthcare-associated infections (HCAIs) including through safe and clean birth practices, hand hygiene, umbilical cord care, and early exclusive breastfeeding - When participants describe maintaining sanitary spaces through appropriate organization of the unit or disinfection of equipment and the clinical environment - When participants describe infection detection through clinical identification, sample and bacterial culture protocols, and effective laboratory-ward communication - When participants describe proper care after infection detection including target anti-infective therapy and supportive care to manage the effects of sepsis | - When participants describe components outside of assurance of clean birth and care settings, early detection of neonatal infections, and appropriate treatment of neonatal infections in settings providing newborn care services - When participants describe factors that make it difficult to maintain infection prevention and control measure at the healthcare facility |
| Infection prevention & control | Infection Prevention and Control (IPC) practices, including the assurance of clean birth and care settings, early detection of neonatal infections, and appropriate treatment of neonatal infections in settings providing newborn care services, are critical for reducing neonatal infections. | Barriers | - When participants describe a need for improvement in the training or implementation for prevention of healthcare-associated infections (HCAIs) including through safe and clean birth practices, hand hygiene, umbilical cord care, and early exclusive breastfeeding - When participants describe inadequate neonatal infection prevention through the organization of the unit or disinfection of equipment and the clinical environment - When participants describe a lack of infection detection through clinical identification, sample and bacterial culture protocols, or effective laboratory-ward communication - When participants describe inadequate care after infection detection including target anti-infective therapy and supportive care to manage multi-body-system effects of sepsis | - When participants describe components outside clean birth and care settings, early detection of neonatal infections, and appropriate treatment of neonatal infections in settings providing newborn care services - When participants describe factors that make it difficult to maintain infection prevention and control measure at the healthcare facility |
| Linkage to maternal care | The linkage of newborn care to maternal care at every level of healthcare delivery and phase of care along the continuum of care from antenatal through intrapartum to postnatal is central to improving newborn survival rates. Additionally, hospitals must ensure smooth connection with special care for high-risk pregnancies (including preterm) and maternal mental health. | Facilitators | - When participants describe easy accessibility of antenatal services including nutritional assessment and interventions, fetal assessment, preventative measures for infections, and treatment of common physiological symptoms associated with pregnancy - When participants describe the effective organization of essential services including reception, triage and assessment area, immediate postnatal high-care area, sick newborn ward, mother care ward, neonatal intensive care units, and theatre for performing caesarean sections are organized so that caregivers and neonates remain in the same care space. - When participants describe respectful maternity care and effective communication with parents throughout each stage of care, particularly for high-risk pregnancies or those requiring specialized care - When participants describe easy accessibility to support services for perinatal and post-natal mental disorders, including after discharge | - When participants describe components outside of assurance of connection between newborn care and maternal care at every level of healthcare delivery or connection with special care for high-risk pregnancies (including preterm) and maternal mental health. - When participants describe factors prevent strong linkage to maternal care |
| Linkage to maternal care | The linkage of newborn care to maternal care at every level of healthcare delivery and phase of care along the continuum of care from antenatal through intrapartum to postnatal is central to improving newborn survival rates. Additionally, hospitals must ensure smooth connection with special care for high-risk pregnancies (including preterm) and maternal mental health. | Barriers | - When participants describe a lack of antenatal service availability or usage - When participants describe the ineffective organization of essential services including reception, triage and assessment area, immediate postnatal high-care area, sick newborn ward, mother care ward, neonatal intensive care units, and theatre for performing caesarean sections are organized preventing the caregivers and neonates from remain in the same care space. - When participants describe factors that prevent respectful maternity care and hinder communication with parents throughout each stage of care, particularly for high-risk pregnancies or those requiring specialized care - When participants describe factors making support services for perinatal and post-natal mental disorders, including after discharge difficult to access | - When participants describe components outside of assurance of smooth connection between newborn care and maternal care at every level of healthcare delivery or connection with special care for high-risk pregnancies (including preterm) and maternal mental health. - When participants describe factors support and enable strong linkage to maternal care |
| Post-discharge follow-up | Continuity of care between the facility and community-based services, along with ensuring good health practices continue at home improves newborn outcomes after discharge. It also ensures that any problems related to growth and development may be identified and treated early. | Facilitators | - When participants describe strategies for post-discharge assessment of aspects including feeding and growth, general health, immunizations, sensory ability, and motor development - When participants describe effective parental education on topics including nutrition, medication, danger signs, safe sleeping, etc. - When participants describe methods for identification and follow-up screening with newborns at risk for neuro-developmental disability - When participants describe available services families that require on-going treatment plans | - When participants describe components outside of assurance of smooth connection between newborn care and maternal care at every level of healthcare delivery or connection with special care for high-risk pregnancies (including preterm) and maternal mental health. - When participants describe factors related to post-discharge care that are difficult to access and prevent high quality SSNC from occurring |
| Post-discharge follow-up | Continuity of care between the facility and community-based services, along with ensuring good health practices continue at home improves newborn outcomes after discharge. It also ensures that any problems related to growth and development may be identified and treated early. | Barriers | - When participants describe a lack of post-discharge assessment of aspects including feeding and growth, general health, immunizations, sensory ability, and motor development - When participants describe ineffective or unavailable parental education on topics including nutrition, medication, danger signs, safe sleeping, etc. - When participants describe insufficient methods for identification and follow-up screening with newborns at risk for neuro-developmental disability - When participants describe a lack of services families that require on-going treatment plans | - When participants describe components outside of assurance of smooth connection between newborn care and maternal care at every level of healthcare delivery or connection with special care for high-risk pregnancies (including preterm) and maternal mental health. - When participants describe factors related to post-discharge care that are easy to access and enable high quality SSNC |
| Financing | Securing funding for small and sick newborn care will require assessment of current investments and needs based on economic evidence, formation of an investment case based on the evidence, and pooling of resources to ensure financial protection of patient families. | Facilitators | - When participants describe strategies for generating economic evidence for investing in high-quality care for small and sick newborns through cost-assessment for setup, running, and maintenance - When participants describe strategies to develop an investment case based on economic evidence by assessing the investment required to reach targets, the potential return on investment and existing gaps in coverage and financing. - When participants describe strategies for mobilization of additional resources to minimize out-of-pocket payments | - When participants describe components outside of assessment of current investments and needs based on economic evidence, formation of an investment case, ensuring financial protection of patient families. - When participants describe factors related to the assessment of current investments and needs based on economic evidence, formation of an investment case that make it harder to provide SSNC and don’t ensure the financial protection of patient families |
| Financing | Securing funding for small and sick newborn care will require assessment of current investments and needs based on economic evidence, formation of an investment case based on the evidence, and pooling of resources to ensure financial protection of patient families. | Barriers | - When participants describe insufficient efforts to generate economic evidence for investing in high-quality care for small and sick newborns through cost-assessment for setup, running, and maintenance - When participants describe insufficient efforts to develop an investment case based on economic evidence by assessing the investment required to reach targets, the potential return on investment and existing gaps in coverage and financing. - When participants describe insufficient or ineffective strategies for mobilization of additional resources to minimize out-of-pocket payments | - When participants describe components outside of assessment of current investments and needs based on economic evidence, formation of an investment case, ensuring financial protection of patient families. - When participants describe factors related to the assessment of current investments and needs based on economic evidence, formation of an investment case that make it easier to provide SSNC and ensure the financial protection of patient families |

https://newborntoolkit.org/toolkit/leadership-and-governance/overview
