## Supplemental table 1 for "From warehouse to ward: Applying implementation research methods to the device identification, qualification, distribution, and management process within the NEST360 alliance"

**Supplemental Table 1.** Inventory of strategies used to improve the implementation of device qualification, management, and distribution activities in NEST360

| **Strategies** | **Strategy Explanation, Adapted from ERIC Definitions** | **ERIC Strategy** |
| --- | --- | --- |
| Target product profiles (TPPs) (A) | NEST360 developed TPPs which define the ideal specifications, usability, affordability, and sustainability criteria for SSNC devise to ensure they are effective, durable, and suitable for low-resource settings. | Create or change credentialing and/or licensing standards |
| Device qualification process (B) | Medical devices for SSNC were rigorously evaluated against TPP criteria, tested for performance under challenging conditions (e.g., heat, dust, power surges), and assessed for safety, durability, affordability, and suitability before receiving NEST360 approval. Qualification process occurred in the United States and Malawi. | Change physical structure and equipment/tailor strategies |
| Pre-installation health facility assessment (C) | NEST360 conducted a pre-installation assessment that evaluates the infrastructure, resources, and readiness of facilities to ensure they can support and sustain SSNC devices. | Assess for readiness and identify barriers and facilitators |
| Develop and distribute educational material (D) | NEST360 created technical modules, job aids, and practical scenarios to enhance BMET training and facilitate effective knowledge acquisition. | Develop educational material, distribute educational material |
| Preservice training (E) | NEST collaborated with educational institutions to equip local BMET programs with SSNC devices, enabling students to develop maintenance skills before graduation. | Work with educational institutions |
| Mentorship (F) | NEST360 BMETs offered regular mentorship to facility-level BMETs, both in person and virtually, to assist them in repairing nonfunctioning devices. | Conduct ongoing training |
| Conduct educational (quality improvement (QI) meetings (G) | METs actively participated in facility-level quality improvement meetings to review change ideas and data, and they also led BMET-specific QI projects that contributed to enhancing care quality. | Conduct educational meetings |
| Conduct ongoing training (BMETs) (H) | NEST360 conducted ongoing training initiatives, including country-level mentorship, facility-level coaching, and peer-to-peer learning meetings. | Conduct ongoing training |
| Capture and share local knowledge (I) | The NEST360 team analyzed routinely collected BMET data to identify areas for improvement and highlight successes to share with healthcare facilities. | Capture and share local knowledge |
| Identify and prepare champions (J) | BMET ward improvement team members were appointed to foster collaboration and actively involve BMETs in QI activities. | Identify and prepare champions |
| NEST country-level BMETs (K) | Each country had a NEST360-appointed BMET who provided training, mentorship, and support to facility-level BMETs. | Provide local technical assistance |
| Hatch (L) | Hatch Technologies, a nonprofit organization, was initially established to support NEST360’s device distribution, including sourcing and shipping needs, but has since expanded to provide commercial support through authorized distributors. | Provide local technical assistance |
| Hadli (M) | A remote device monitoring system that reports data on device utilization and facility power quality to help BMETs maintain devices. | Develop and implement tools for quality monitoring |
| NEST-IT (N) | NEST360 created an electronic dashboard that provides data views at the facility, country, and Alliance levels. This dashboard visualizes an interdisciplinary set of indicators across various levels of aggregation, supporting the diverse efforts of stakeholders working to improve neonatal care. It enables stakeholders to monitor clinical processes and outcomes effectively. | Develop and organize quality monitoring systems |
| Audit and provide feedback (O) | Country-level BMETs conducted quarterly tracking and reporting of activities to summarize performance, enabling facility and country leaders to monitor progress, evaluate outcomes, and implement necessary adjustments. | Audit and feedback |
| Providing tools (P) | NEST360 equipped each facility with a toolbox to enable BMETs to carry out both preventive and corrective maintenance effectively. | Change physical structure and equipment |
| Providing consumables and spare parts (Q) | NEST360 supplied facilities with the necessary consumables and spare parts to maintain devices at no cost. | Change physical structure and equipment |
| Providing device bundles (R) | During installation, all NEST360-supported facilities were provided with a comprehensive package containing all the necessary devices to deliver SSNC effectively. | Mandate change |
| Collaborate with stakeholders (MoH, facilities) (S) | NEST360 engaged a diverse group of stakeholders—including Ministry of Health officials, facility leadership, professional organizations, clinicians (maternal and neonatal), and BMETs—to raise awareness about the critical role of BMETs, emphasize the importance of maintaining buffer stock at the national level, and initiate discussions on long-term sustainability. | Involve executive boards |
| Access new funds (national and global) (T) | The NEST360 program engaged with various funding organizations and collaborated with Ministries of Health to secure long-term financial support for NEST360 activities, including training programs and the ongoing procurement of essential items such as consumables and spare parts. | Access new funding |
| Develop a BMET model maintenance workshop (U) | A fully equipped model workshop was established in each country where NEST360 operates. These workshops were designed with sufficient space, resources, and tools to serve as a blueprint for national leadership, showcasing the standards that all healthcare facilities should meet to support effective BMET activities. | Model and simulate change and change physical structure and equipment |

*Note. The letters after each strategy correspond to the letters used in the IRLM to map strategies to contextual factors and mechanisms of action.*

*Biomedical engineers and technicians (BMETs); NEST-Implementation tracker (NEST-IT); Ministry of Health*
