## Supplemental table 2 for "From warehouse to ward: Applying implementation research methods to the device identification, qualification, distribution, and management process within the NEST360 alliance"

**Supplemental Table 2.** Detailed Definitions of NEST360’s Device Identification, Qualification, Management, and Distribution Implementation Outcomes

| **Dimension** | **Measurement** | **Time Period** | **Data Source** | **Outcome** |
| --- | --- | --- | --- | --- |
| **Reach** (Distribution) | Number of facilities that received technology from NEST360 | By end of phase 1 (2023) | Routine programmatic data (NEST-IT) | **66 facilities** received NEST360 qualified devices  Malawi-37 units, Kenya-13 units,Tanzania-7 units, Nigeria-11 units |
| **Reach** (Qualification) | Number of devices that have received NEST360 qualification | By end of Phase 1 (2023) | Qualitative interview | **29 devices were approved** through the NEST360 qualification process­­­­­ |
| **Reach** (Qualification) | Number and % of categories that have at least one qualified device approved of the 15 product categories | By end of Phase 1 (2023) | Qualitative interview | **12 of the 15 (87%) proposed SSNC categories** have at least one qualified device approved |
| **Effectiveness** | Categorical determination of if there were enough CPAP devices to cover all eligible babies | By end of Phase 1(for most facilities this is Q4 of 2023, but some is Q3 of 2023) | Routine programmatic data (NEST-IT) | **Malawi: Enough:70.3% (26/37 units)**  Not enough </=10 days: 21.6% (8/37)  Not enough > 10 days:5.4% (2/37)  Missing data: 1 unit*  **Kenya: Enough: 69.2% (9/13 of units)**  Not enough </=10 days: 15.4% (2/13)  Not enough > 10 days: 15.4% (2/13)  **Tanzania**: **Enough:100 % (7/7 of units)**  Not enough </=10 days: 0% (0/7)  Not enough > 10 days: 0% (0/7)  **Nigeria**: **Enough: 81.8% (9/11 of units)**  Not enough </=10 days: 9.1% (1/11)  Not enough > 10 days: 0% (0/10)  Missing data: 1 unit*  **NEST program: Enough: 75% (51/68 of units)**  Not enough </=10 days: 16.2% (11/68)  Not enough > 10 days: 5.9% (4/68)  Missing data: 2 units*  *Data on CPAP device availability was missing from one site in Nigeria and one in Malawi |
| **Effectiveness** | Categorical determination of if there were enough phototherapy devices to cover all eligible babies | By end of Phase 1(for most facilities this is Q4 of 2023, but some is Q3 of 2023) | Routine programmatic data (NEST-IT) | **Malawi: Enough: 81.1% (30/37 units)**  Not enough </=10 days: 10.8% (4/37)  Not enough > 10 days: 2.7% (1/37)  Missing data: 2 units*  **Kenya: Enough: 61.5% (8/13 of units)**  Not enough </=10 days: 30.8% (4/13)  Not enough > 10 days: 7.7% (1/13)  **Tanzania: Enough: 57.1% (4/7 of units)**  Not enough </=10 days: 42.9% (3/7)  Not enough > 10 days: 0% (0/7)  **Nigeria: Enough: 90.9% (10/11 of units)**  Not enough </=10 days: 9.1% (1/11)  Not enough > 10 days: 0% (0/10)  **NEST program: Enough: 76.5% (52/68 of units)**  Not enough </=10 days: 17.6% (12/68)  Not enough > 10 days: 2.9% (2/68)  Missing data: 2 unit*  *Data on Phototherapy device availability was missing from two sites in Malawi |
| **Effectiveness** | Percentage of newborn care devices that were functional, includes those provided by NEST360 and those sourced through existing channels | Jan-Dec 2024 | Hatch ((Asset and ticketing tracking) | **87%** **of all newborn care devices were functional,** including those provided by NEST360 and those sourced through existing channels |
| **Adoption** (of NEST qualified devices) | Number of NEST360 qualified devices installed at NEST360 facilities | By end of Phase 1 (2023) | Hatch ((Asset and ticketing tracking) | **2677 devices installed** at NEST360 facilities |
| **Implementation Acceptability** (of the device implementation strategies) | Responses from PO4/PO5, HATCH, and BMETs on acceptability of the identification, qualification, distribution, and management process | From start to interview date (May-July 2024) | Qual interviews | Stanford Lighting Reports |
| **Implementation Adaptations** (to the implementation strategies) | Changes that occurred to the device identification, qualification, distribution and management process throughout Phase 1 | By end of Phase 1 (2023) | Qual interviews | See Stanford Lighting Report (ex. developing a qualification site in Malawi to promote sustainability) |
| **Implementation Feasibility** (having the necessary personnel present) | Does the facility have a BMET available during day or night shift? (does not have to be newborn ward specific) | End of Phase 1 (for most facilities this is Q4 of 2023, but some is Q3 of 2023) | Routine programmatic data (NEST-IT) | **Malawi: 86% (32/37 units) ***  **Kenya: 100% (13/13 of units)**  **Tanzania: 100% (7/7 of units)**  **Nigeria: 100% (11/11 of units)**  **NEST program: 93% (63/68 of units)***  Missing data: 5 units from Malawi  *Data on BMETs was missing from 5 units in Malawi because of disruptions from Hurricane Freddy |
| **Implementation Feasibility** (to the distribution of devices) | Are distributors able to get the qualified devices into the countries and installed at the individual facilities | By end of Phase 1 (2023) | Qual interviews | See Stanford Lighting Report and device distributors table |
| **Implementation Fidelity** (to the maintenance recommendations) | Percentage of facilities performing monthly PPM for 5 of the 6 reported categories (glucometer, pulse ox, CPAP, O2 concentrator, radiant warmer, phototherapy) | Feb-Dec 2022 vs. Jan-Dec 2023 | Routine programmatic data (NEST-IT) | **58.6% in 2022**  **70.8% in 2023**  * Some facilities were missing from each month, so 58.6% and 70.8% is reflective of the average percentage of facilities performing PPM for 5/6 months of those who provided any data related to PPM during the months of Jan-June 2024 |
| **Implementation Fidelity** (to qualification) | Did all NEST360 qualified devices go through the full identification and qualification process | By end of Phase 1 (2023) | Qual interviews | **Yes**, of the 29 devices approved all went through the full qualification process  Hatch does sell products that are not NEST-360 qualified however they are not marketed as NEST-qualified |
| **Maintenance** (Continuing to follow recommendations for keeping devices functional after implementation efforts end) | Percentage of facilities performing monthly PPM for 5 of the 6 reported categories (glucometer, pulse ox, CPAP, O2 concentrator, radiant warmer, phototherapy) | Jan-June 2024 | Routine programmatic data (NEST-IT) | **81.9%**  *Some facilities were missing from each month, so 81.9% is reflective of the average percentage of facilities performing PPM for 5/6 months of those who provided any data related to PPM during the months of Jan-June 2024 |
| **Maintenance** (Device qualification, distribution, and managements process) | Areas where the device teams have already, are beginning to, or should consider focusing their attention to ensure sustainability of their work | By time of interviews (May-July 2024) | Routine programmatic data (NEST-IT) | See Stanford Lighting Report |
